# Lymphocyte Stability Following Immune Checkpoint Blockade Is Coupled to Mitochondrial Haplogroup and Clinical Outcome in Advanced Melanoma

**DOI:** 10.64898/2026.10.30.26366523

**Authors:** Shawn Sun, Maksim C.F. Chan, Robert A. Watson, Sophie MacKay, Martin Little, Orion Tong, Andrea Farkas, Guangyi Niu, Lea R.Z. Cohen, Gusztav Milotay, Dylan Muldoon, Nicholas Coupe, Miranda J. Payne, Brian Shine, Mark R. Middleton, Paul Klenerman, Benjamin P. Fairfax

**Author notes:** These authors contributed equally (Shawn Sun, Maksim C.F. Chan).

## Abstract

Early biomarkers of durable response to immune checkpoint blockade (ICB) are urgently needed. We identify lymphocyte stability (LS) – preservation of peripheral lymphocyte counts after one cycle of ICB – as an on-treatment predictor of overall survival (OS) in metastatic melanoma (MM): patients with unstable lymphocyte counts (LS-U; post-ICB/baseline ≤0.8) have markedly increased risk of death. Metabolic profiling of patient T cells demonstrates ICB enhancement of mitochondrial bioenergetics, whilst CD8+ T cells from LS-U patients mount an exaggerated transcriptional response to ICB incorporating upregulation of mitochondrial pathways. Given these findings we assessed the relationship of LS with mitochondrial haplogroup, revealing carriage of haplogroup T (HG-T), present in ∼10% of white British individuals, is associated with both LS-U and impaired OS, with mediation analysis implicating LS mediation. HG-T carriers have higher CD8+ T-cell pre-treatment mitochondrially encoded transcript abundance and, in paired scRNA-seq data, fewer progenitor-exhausted cells, ICB hyperresponsivity, apoptotic program induction and reduced clonal persistence – consistent with baseline mitochondrial priming. These findings position LS as a clinically accessible marker of ICB outcome linked to inherited mitochondrial variation.

**Highlights:**

- Lymphocyte stability is a pan-cancer predictor of survival and durable response following immune checkpoint blockade
- ICB elicits coordinated mitochondrial transcriptional and functional responses in T cells
- LS-U mediates the link between mitochondrial HG-T and poor overall survival following checkpoint immunotherapy
- HG-T CD8+ T cells are transcriptionally primed at baseline but show enhanced apoptosis and reduced clonal persistence after ICB

## Introduction

Immune checkpoint blockade (ICB), consisting of monoclonal antibodies that target the checkpoint proteins PD-1, CTLA-4 and LAG-3, forms the mainstay of treatment for patients with advanced unresectable or metastatic melanoma (MM) and is increasingly central to the management of many other cancers. Marked heterogeneity between patients in terms of clinical benefit from ICB is apparent however, with over half of patients treated with combination anti-PD-1 (nivolumab) and anti-CTLA-4 (ipilimumab) for MM having disease-specific survival at 10 years, the rest progressing and dying within five years^1^. A comprehensive understanding of the determinants of variation in response to ICB promises to advance treatment stratification and development and is of major clinical and immunological interest.

Although early efforts to explain this heterogeneity focused on tumour features that confer sensitivity or resistance to ICB, it is increasingly clear that patient systemic immunity is cardinal to anti-tumour activity. One such factor is the ability to mount a robust lymphopoietic response after ICB initiation. This was originally described in early clinical trials of ipilimumab for MM, where patients who maintained peripheral blood lymphocyte counts (LC) after two cycles of treatment had a reduced risk of death^2^. Subsequent small retrospective studies linked on-treatment lymphopenia to poorer oncological outcomes of ICB in melanoma and lung cancer^3,4^. Analysis of large datasets revealed a subtle fall in lymphocyte count is typical between the first and second cycles of anti-PD-1 based treatment for cancer^5,6^. We hypothesise that the lymphopoietic response to ICB represents effective re-invigoration of T-cell function, linked to T-cell clonal expansion and antigen recognition, together with parameters of overall immune fitness, including sufficient lymphotropic cytokines. In keeping with this, human genetic studies implicate IL-7, a cytokine central to lymphogenesis and peripheral T-cell homeostasis^7^, in mediating the lymphopoietic response to ICB. Carriers of an intronic polymorphism in *IL7,* associated with a cryptic splice-variant and increased *IL7* expression, show a reduced ICB-induced LC fall^5,6^. IL-7 is pleiotropic, antagonising the intrinsic mitochondrial apoptotic pathway through JAK-STAT5-induced BCL2 expression and promoting memory T-cell differentiation^8,9^. Other crucial cytokines include IL-2 for T-cell expansion, and IL-15 for T-cell survival. Beyond lymphotropic factors, clinical benefit from ICB is linked to a sufficient population of stem-like progenitor exhausted T cells (TPEX), which mediate the proliferative response to ICB^10–13^. Other favourable peripheral factors in patients with MM include greater diversity of expanded large CD8+ T-cell clones, with increased expression of genes linked to T cell receptor signalling and mitochondrial translation being observed in CD8+ T cells of patients with durable responses^14^. Notably, mitochondrial function is exquisitely linked to T-cell state, with dynamic changes in mitochondrial morphology being linked to T-cell activation^15^. In CAR-T cell therapy, mitochondrial fitness is tightly associated with cellular persistence, memory differentiation and resistance to exhaustion^16,17^, while effective T-cell infiltration and sustained functionality within tumours are critically entwined with mitochondrial metabolic state and oxidative fitness^18,19^.

Previously, in a prospective cohort of patients with MM treated with anti-PD-1 based singlet or combination ICB, we observed that maintenance of LC after treatment, termed lymphocyte stability (LS), was associated with both progression-free survival (PFS) and overall survival (OS)^6^. Here, we have dissected the underpinnings of LS in the Oxford Cancer Immunotherapy Toxicity and Efficacy cohort (OxCITE), comprising 417 prospectively recruited patients receiving ICB for cancer with pre- and on-treatment LC available (Table S1, Extended Figure 1). After replicating our original observation of impaired OS in patients with MM with a post-ICB LC ≤0.8 baseline (lymphocyte-unstable, LS-U), we defined the core transcriptional determinants of LS in CD8+ T cells. Patients with post-ICB LC >0.8 baseline LC (lymphocyte-stable, LS-S) displayed transcriptional induction of IL-7 signalling and mitochondrial biogenesis, whereas those with LS-U demonstrated pronounced induction of pathways linked to oxidative phosphorylation and mitochondrial activity. Functional analysis of patient-derived primary T cells pre- and post-ICB demonstrates pervasive enhancement of T-cell metabolic parameters by ICB, consistent with the crucial role of T-cell mitochondria in ICB response. We reasoned that if mitochondrial function contributes causally to LS, a relationship between germline mitochondrial variation and LS may exist. To test this, mitochondrial haplogroups were imputed across patients from germline genotyping. This revealed a striking association between LS and carriage of mitochondrial haplogroup T (HG-T), which encodes non-synonymous HG-specific Complex I variants and is carried by approximately 10% of the white British population^20^. HG-T carriers are more likely to be LS-U and have reduced OS, with mediation analysis supporting a causal relationship between HG-T effect on LS and association with OS. Consistent with the haplogroup genetic divergence, HG-T carriers have altered expression of a discrete subset of mitochondrial encoded genes core to respiratory complexes. Single-cell RNA-seq analysis of CD8+ T cells further linked HG-T to reduced proportions of progenitor exhausted cells. Our findings demonstrate that LS forms a robust and easily discernible on-treatment predictive biomarker of survival post-ICB in MM, and HG-T carriage is one determinant of impaired LS. This work underlines the dependence of durable response to ICB on systemic immune fitness, and highlights T-cell mitochondrial function as a promising therapeutic target in cancer immunotherapy.

## Results

### Lymphocyte stability as a pan-cancer predictor of post-ICB survival

#### Validation of LS as a predictor of post-ICB survival

We first sought to replicate our original observation linking LS to survival^6^ using further internal and external validation cohorts (Figure 1A). In a newly-recruited OxCITE validation cohort consisting of 168 MM patients with a median follow-up of 41.6 months (95% CI 32.7-48.3), LS-U patients were again observed to have worse overall survival (OS) compared to LS-S patients (Figure 1B; log-rank P = 1.1×10^-3^), with LS-U status having a hazard ratio (HR) of 2.26 for death (95% CI 1.37-3.73), and reduced 12-month median survival (57.6% vs 82.8%; LS-U median OS: 12.8 months (95% CI 7.6-NR) vs. LS-S median OS: 63.7 months (95% CI 44.2-NR)). This finding was further validated using an independent external cohort of 170 MM patients not incorporated into OxCITE (Table S2), with median follow-up of 58.7 months from initiating ICB (95% CI 46.1-78.3). Again, patients with LS-U demonstrated worse OS than LS-S patients (Figure 1C, log-rank P = 2.7×10^-4^; LS-U median OS: 13.8 months (95% CI 5.9-24.7) vs. LS-S median OS: 28.7 months (95% CI 19.4-NR)) and a HR of 2.18 for death (95% CI 1.42-3.35), mirroring the OxCITE cohort results. Similar findings were observed for progression-free survival (PFS) in both cohorts (Figures S1A-B), as well as after incorporating the non-melanoma cancer patients in the OxCITE cohort (Figure S1C, n=323; OS log-rank P < 1×10^-4^, HR = 2.39, 95% CI 1.7-3.38), highlighting LS as a potential pan-cancer biomarker of ICB response.

**Figure 1:**
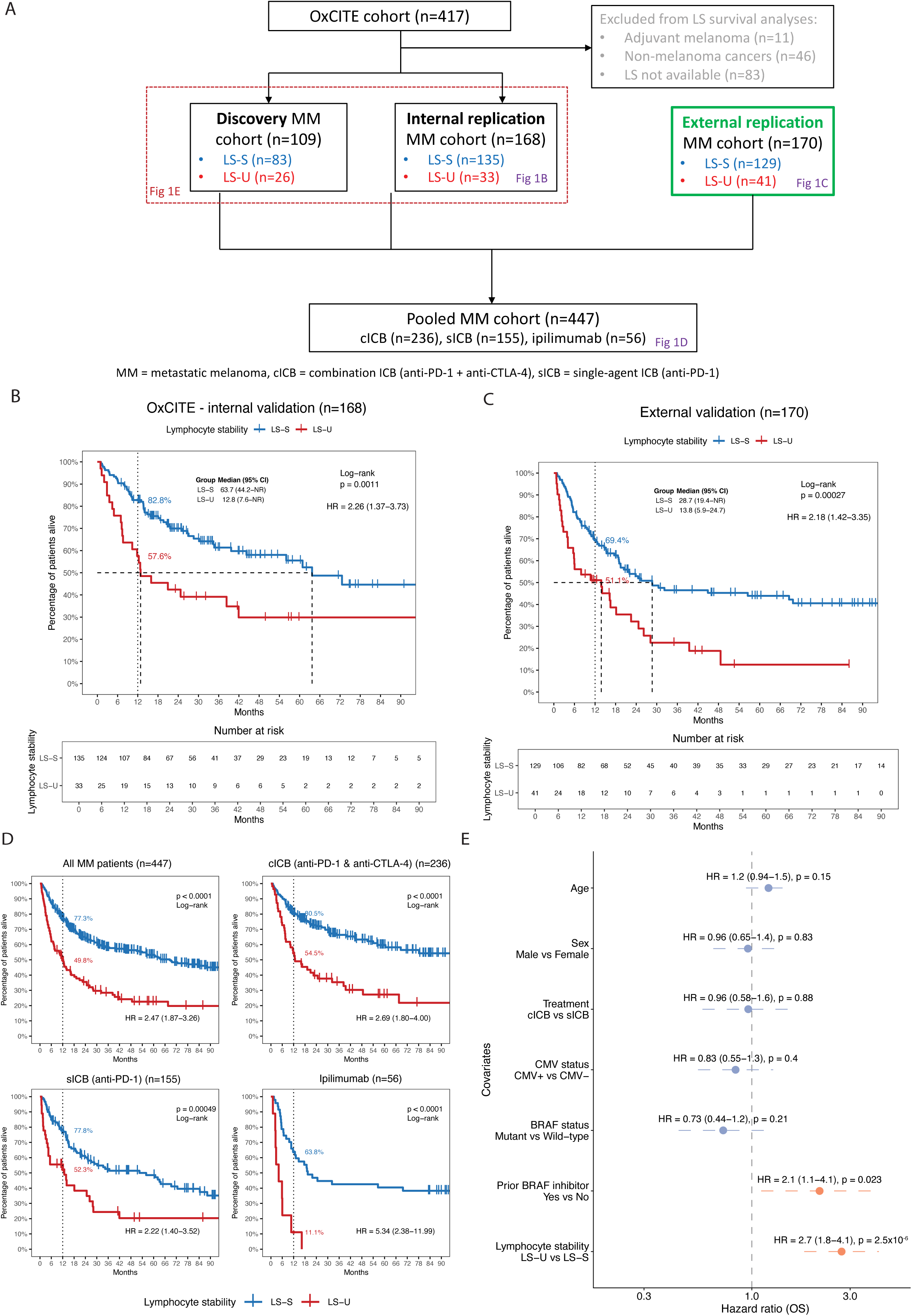
Lymphocyte stability (LS) is an on-treatment predictor of survival after immune checkpoint blockade. **(A)** Patient allocation across discovery, internal replication, external replication and pooled metastatic cohorts. **(B-C)** Kaplan–Meier overall survival (OS) analyses in the OxCITE internal (n=168) and external (n=170) melanoma validation cohorts. **(D)** OS in pooled melanoma patients (n=447), stratified by ICB regimen (combination ICB, anti-PD-1 or anti-CTLA-4). **(E)** Multivariable Cox regression of OS adjusted for age, sex, regimen, CMV serostatus, BRAF status, prior BRAF inhibitor exposure and LS.

To assess the robustness of LS across treatment regimens, data were combined across cohorts (n=447), splitting the survival analyses by treatment type. This showed the association between LS and survival to be robust across all regimens, with LS-U patients demonstrating poorer OS, with a 2.22 HR for death in recipients of anti-PD-1 alone (n=155, P = 4.9×10^-4^), rising to 2.69 HR amongst combination ICB (anti-PD-1 + anti-CTLA-4) recipients (n=236, P < 1×10^-4^), and a 5.34 HR in recipients of anti-CTLA-4 alone (n=56, P < 1×10^-4^) (Figure 1D).

Throughout these analyses, patients were classified as LS-S or LS-U using a post-treatment (C2) to pre-treatment (C1) lymphocyte count ratio cutoff at 0.8 as defined previously^6^. This cutoff provided the maximal log-rank statistics in the OxCITE melanoma cohort and after merging with the external validation cohort (Figure S1D). Time-dependent receiver operating characteristic (ROC) analyses across cohorts indicated the optimal cutoff ranged between 0.66-0.92 (Figure S1E). Given this, a threshold of 0.8 for LS-U was retained for all downstream analyses.

#### Interaction between LS and other prognostic indicators

In keeping with LS providing independent prognostic information, major clinical characteristics were balanced between LS-S and LS-U patients (Extended Table 1). To assess whether LS provides additional information to characterised prognostically informative blood-based biomarkers, the relationship between LS and pre-treatment neutrophil-to-lymphocyte ratio (NLR)^21^, platelet-to-lymphocyte ratio (PLR)^22^ and lymphocyte-to-monocyte ratio (LMR)^23^, was assessed across patient samples within OxCITE, finding no difference in these biomarkers between LS-S and LS-U groups (Figure S2A). However, subgroup analysis demonstrated that among patients with above median NLR – a prognostically unfavourable marker – LS-U patients had a 5-fold higher risk of death compared to LS-S patients (Figure S2B, n=234; NLR-high HR=5.07, 95% CI 2.91-8.84, P < 1×10^-3^). The relationship between LS and CMV serostatus, strongly associated with lymphocyte count and previously found to impact anti-PD-1 treatment response in patients with MM^24^, was also assessed. Despite CMV-seropositive patients having higher baseline and on-treatment lymphocyte counts, the C2:C1 ratio did not significantly differ between CMV groups (Figure S2C). Again, this analysis showed LS stratified post-ICB survival, with LS-U patients having greater than two-fold risk of death irrespective of CMV serostatus (Figure S2D, n=250; CMV-seropositive HR=2.76, 95% CI 1.36-5.6, P = 3×10^-3^; CMV-seronegative HR=2.45, 95% CI 1.48-4.05, P < 1×10^-3^). Similar observations were made when controlling for other potential confounding covariates with a Cox proportional hazard model; adjusting for age, sex, treatment regimen, CMV serostatus, melanoma *BRAF* mutational status, and prior treatment with BRAF inhibitors. This analysis again demonstrated LS-U patients have an HR of 2.7 for death (Figure 1E, 95% CI 1.8-4.1, P = 2.5 x 10^-6^). Thus, LS provides early treatment response related prognostic information, distinct to that of other pre-treatment blood-based biomarkers, with particular utility in differentiating potentially ICB responsive high-risk patients.

### ICB induction of CD8+ T cell mitochondrial metabolic programs

#### CD8 T cell transcriptomic correlates of LS

Given the reduced clinical benefit of ICB in LS-U patients, we reasoned that T cells from these patients might demonstrate intrinsic resistance to anti-PD-1 based ICB effects. ICB leads to widespread transcriptional re-modelling, particularly within the CD8 T-cell compartment, and this hypothesis was addressed by assessing induced transcriptomic responses according to LS status. Pairwise pre-ICB (C1) and post-ICB (C2) gene expression profiles in sorted CD8+ T cells from 147 MM patients (122 LS-S, 25 LS-U) were compared. To adjust for differences in sample sizes between the LS groups, paired differential expression (DE) analysis was restricted to 10 patients per group and performed across 100 iterations. In contrast to our original assumption, this analysis indicated that the LS-U group consistently displayed more differentially expressed genes in response to ICB than the LS-S group (Figure 2A and Table S3; median 2056.5 vs 582.5 differentially expressed genes [DEGs]; P < 2.2 x 10^-16^). Comparing the effect sizes from all genes with log_2_ fold change estimates computed in ≥80% of iterations, while both groups showed highly concordant transcriptional changes, CD8+ T cells from LS-U patients had a greater median log_2_ fold change in expression (Figure S3A; slope of log_2_FC LS-U vs. log_2_FC LS-S 1.419, 95% CI 1.409-1.43, R^2^ = 0.815, Wald t-test P < 1×10^-3^). Analysis of the top 5000 genes in each group ranked by response magnitude (absolute log_2_FC) further demonstrated the exaggerated transcriptional response to ICB in the LS-U group (Figure S3B). Although there was clear sharing of ICB induced transcripts between groups, we wished to discern any qualitative divergence in DE pathways according to LS status. To maximally leverage the power of all available samples, DE analysis was performed across the full dataset (122 LS-S and 25 LS-U patients, all paired C1 vs C2) identifying 9256 ICB induced DEGs across both datasets (Figures 2C and S3C-D, Table S4). Although more DEGs were detected in the LS-S group here, this was attributable to differences in sample sizes between the datasets. Comparison of DEGs from the full datasets again showed that a markedly greater magnitude of ICB-induced transcriptional remodelling from patients with LS-U was detected (Figure 2B; R^2^ = 0.967, slope = 1.513). Pathway enrichment of DEGs was separately performed according to LS status using Gene Ontology Biological Process (GOBP). This identified 493 significantly enriched pathways in the LS-S group (351 upregulated, 142 downregulated), and 400 in the LS-U group (324 upregulated, 76 downregulated; Table S5, all FDR < 0.05). Notably, ICB caused a strong enrichment of many shared pathways in CD8+ T cells including mitotic DNA replication (GO:1902969), positive regulation of IL-2 production (GO:0032743), and ATP-coupled electron transport (GO:0042773) across both groups (Figure 2C). However, distinct features of mitochondrial function showed differential regulation between groups, with LS-S associated with positive regulation of mitochondrial translation (GO:0070131), calcium ion homeostasis (GO:0051560) and response to IL-7 (GO:0098760). Conversely, CD8+ T cells from LS-U patients demonstrated upregulation of a number of specific oxidative respiratory pathways including positive regulation of cellular respiration (GO:1901857), inner mitochondrial membrane organisation (GO:0007007), as well as pyruvate metabolic processes (GO:0006090).

**Figure 2:**
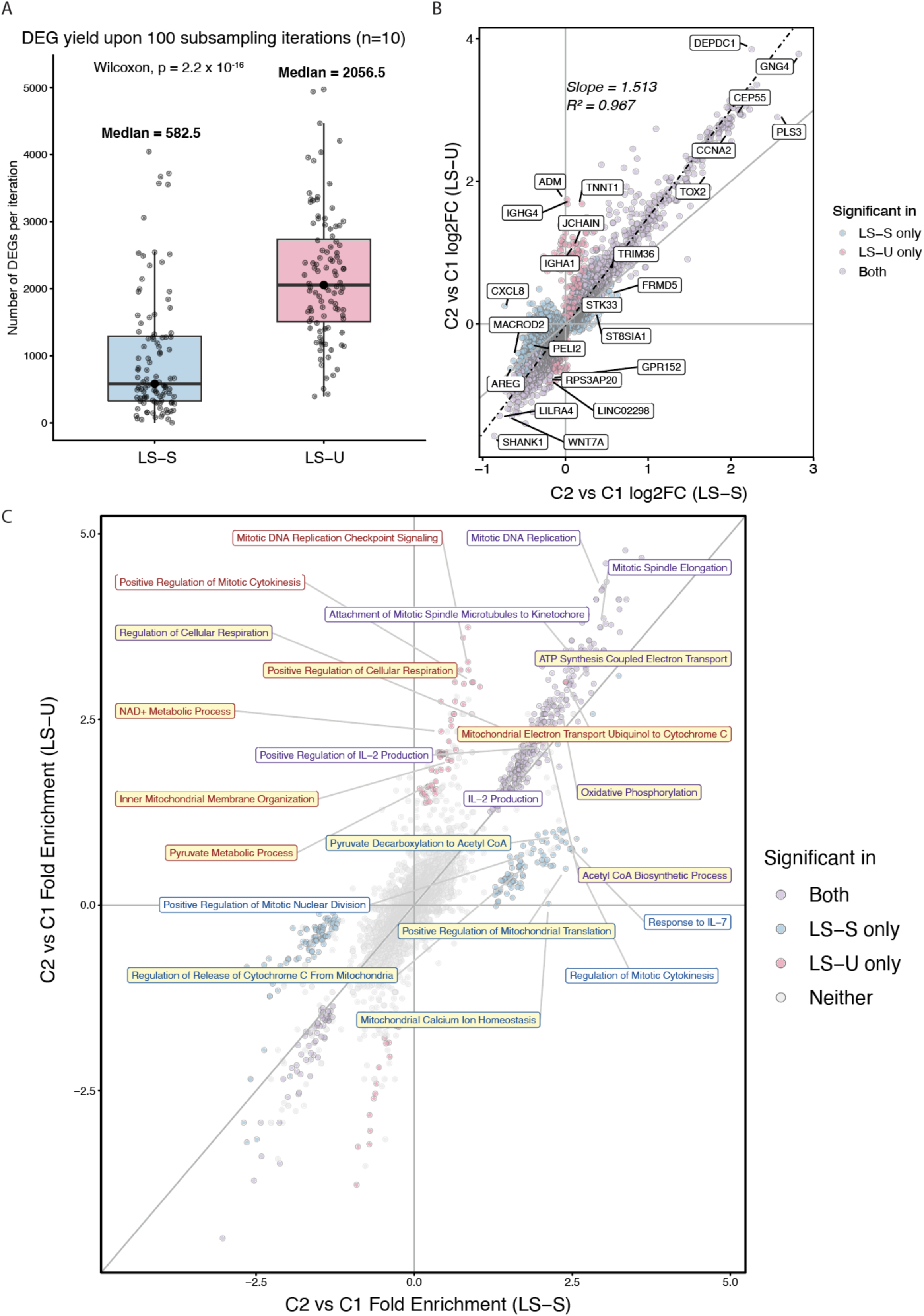
LS-U CD8+ T cells exhibit enhanced ICB-induced transcriptional modelling. **(A)** ICB-induced differentially expressed gene (DEG) counts across 100 subsampling iterations (10 patients per LS group), with median counts and Wilcoxon P value. **(B)** Comparison of ICB-induced gene-level log2 fold changes in LS-U versus LS-S CD8+ T cells, with regression slope and R². **(C)** Directional Gene Ontology Biological Process (GOBP) analysis comparing post-ICB fold enrichment between LS groups. Mitochondrial bioenergetic pathways are highlighted in yellow.

#### Functional analysis links ICB treatment to enhanced T cell bioenergetic demand and capacity

Given the relationship between ICB and induced expression of mitochondrial pathway related transcripts, the functional metabolic correlates of treatment with ICB were explored. To date, the effect of ICB in primary patient T cells on mitochondrial activity has not been functionally assessed. We therefore used the Seahorse XF T-cell Persistence Assay to perform comparative paired analysis of patient sourced T cells pre and post ICB, using negatively selected T cells from patient PBMCs (n=35 paired samples). T-cell oxygen consumption rate (OCR) and extracellular acidification rate (ECAR) were measured, with sequential addition of drugs perturbing mitochondrial electron transport chain function (Oligomycin, BAM15 and rotenone with antimycin A respectively) and bioenergetic profiles assessed throughout (Figure 3A, Table S6).

**Figure 3:**
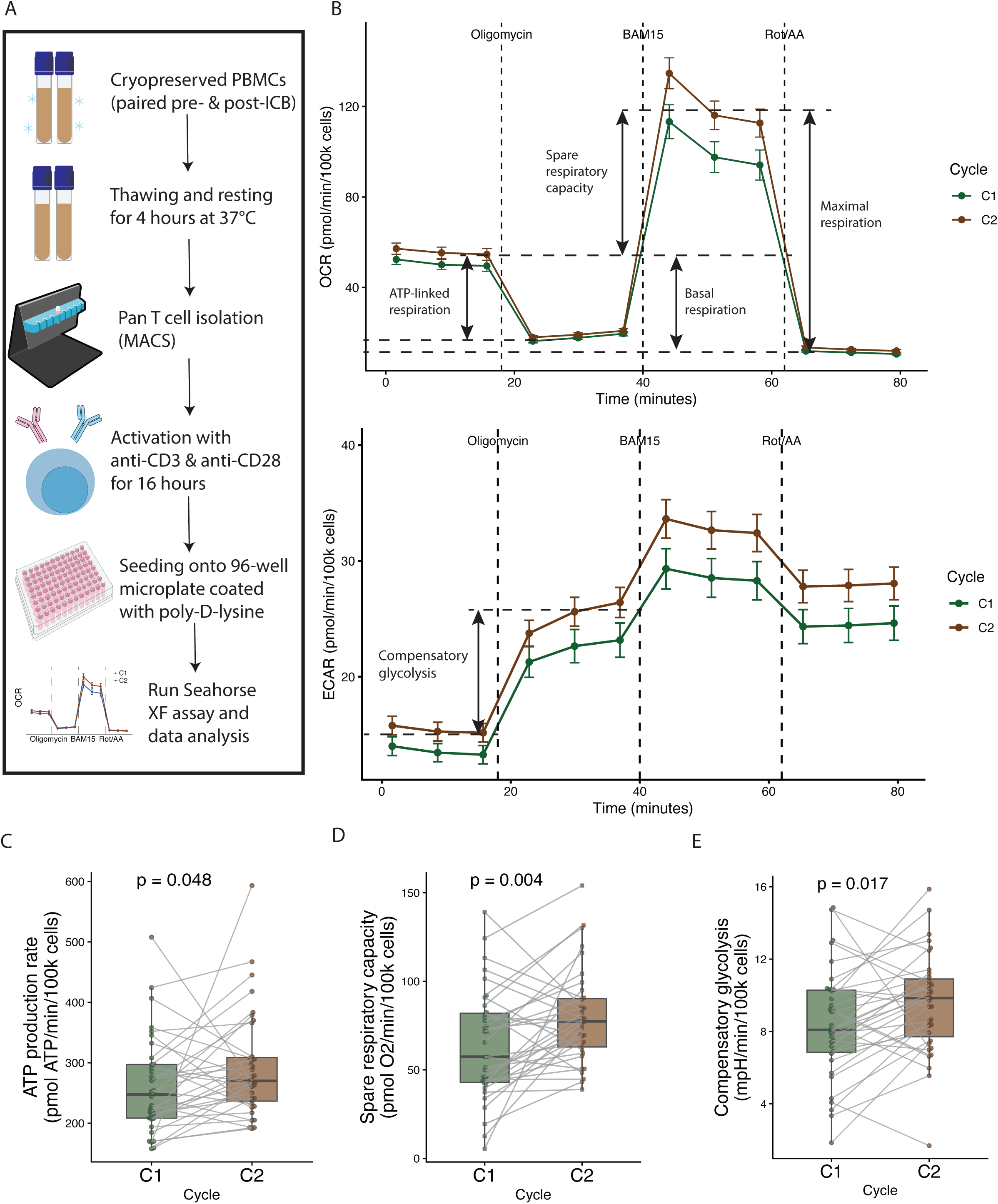
ICB induced T cell metabolic reprogramming. **(A)** Seahorse XF T-cell persistence assay workflow using paired pre- and post-ICB PBMCs. **(B)** Oxygen consumption rate (OCR) and extracellular acidification rate (ECAR) across 12 timepoints including baseline and sequential drug injections. Derived parameters are annotated; values are normalized per 100,000 viable cells. **(C-E)** Paired pre- and post-ICB comparisons of total ATP production, spare respiratory capacity and compensatory glycolysis, respectively.

This demonstrated an ICB induced global increase in T-cell bioenergetic demand, with an upward shift in both oxygen consumption rate (OCR) and extracellular acidification rate (ECAR) kinetic profiles (Figure 3B), from which the metrics ATP-linked respiration, spare respiratory capacity, basal respiration, maximal respiration and compensatory glycolysis could be derived. Post-ICB T-cells had higher total ATP production rates (Figure 3C, P = 0.048), which was driven by coordinated increases in both mitochondrial ATP-linked respiration (Figure S4A, P = 0.044) and glycolytic ATP production (Figure S4B, P = 0.027).

Intriguingly, ICB was observed to enhance T-cell metabolic adaptability, with both increased maximal respiration rate (Figure S4C, P = 0.012) and higher spare respiratory capacity (SRC), both in real terms and also relative to basal metabolic rates (Figure 3D, P = 0.004; Figure S4D, P = 2.6×10^-4^). Notably SRC represents the mitochondrial reserve available to meet acute energetic demands, vital for T-cell survival and function. Upon inhibition of mitochondrial ATP production by oligomycin, post-ICB T cells demonstrated greater compensatory glycolysis, as reflected by upregulated ECAR (Figure 3E, P = 0.017). Together, ICB not only elevates overall energy demand but also enhances metabolic flexibility, enabling T cells to dynamically engage both oxidative and glycolytic pathways in response to stress. Coupled with the distinct hypermetabolic CD8+ T cell state observed in LS-U patients following ICB, these findings implicated mitochondrial function as a potential determinant of variation in lymphocyte stability and ICB outcomes.

### LS serves as a mediator linking Haplogroup T and clinical outcome

#### LS-U is associated with carriage of mitochondrial HG-T

Motivated by the enrichment of mitochondrial metabolic pathways in LS-associated genes, we investigated the potential role for variation in innate mitochondrial characteristics to modulate the interaction between LS and ICB efficacy. Whereas rare deleterious mitochondrial genome variants can lead to heritable mitochondriopathies^25,26^, the relationship between common variation in the mitochondrial genome, secondary to ancestral mitochondrial haplogroup, and mitochondrial function is less well characterised, especially in the context of T-cell immunity. Given the link between LS and mitochondrial function, we reasoned that variation in mitochondrial genetics – where many hundreds of coding and non-coding SNPs differ as maternally inherited, ancestrally shared haplogroups – might directly impact LS. The potential for this to be related to ICB response was further highlighted by a recent association between mitochondrial haplogroups and response to anti-PD-1 in a subset of CheckMate-067 trial participants^27^. Patient samples from the OxCITE cohort were genotyped using the GSA-24v3 array, providing dense mitochondrial genome coverage across 1,060 unique positions. By further imputing mitochondrial SNPs and then predicting haplogroups using Haplogrep 3^28,29^, it was possible to define haplogroups to high accuracy (417 total tested, 414 prediction accuracy of >0.8 and heteroplasmy level >0.9). These could be sub-grouped into 20 haplogroups, with the haplogroups H, U, T, J and K, comprising the majority of the cohort, in proportions consistent with those of the white British population^20^ (Figure S5A). Focusing on haplogroups with a frequency of ≥5% within the OxCITE cohort, the association with the pre- to post-treatment lymphocyte count ratio was assessed. Strikingly, whilst the lymphocyte ratio was comparable amongst H, U, J and K groups, it was notably reduced in HG-T patients when comparing across all OxCITE patients (Kruskal−Wallis P = 0.036, Figure 4A), or just those with MM (Kruskal−Wallis P = 0.018, Figure S5B). Consistently, direct comparison of HG-T with all other Non-T haplogroups confirmed lower lymphocyte ratios in HG-T carriers (Wilcoxon P = 4.6×10^-4^ for MM patients, P = 1.4×10^-3^ for all OxCITE patients; Figures 4B and S5C). The addition of anti-CTLA-4 to anti-PD-1 therapy leads to substantial increases in transcriptional activity relating to metabolic demand and T cell proliferation over anti-PD-1 monotherapy^14,30,31^. In keeping with a relationship between metabolic demand and LS, the association between HG-T carriage and reduced LS was markedly stronger when analysis was restricted to recipients of cICB (Figure S5D). The link between HG-T carriage and LS was further assessed using a linear model across all samples, accounting for potential confounding factors including age, sex, cancer type, treatment regimen and CMV serostatus. This approach again demonstrated an association between LS-U and HG-T carriage (Figure 4C). To ensure that this observation was not being driven by outlier samples, bootstrap analysis was performed across 1,000 iterations, restricting analysis to a 70% subsample of the cohort which further demonstrated >93% of iterations yielded a similar significant association (Figure S5E).

**Figure 4:**
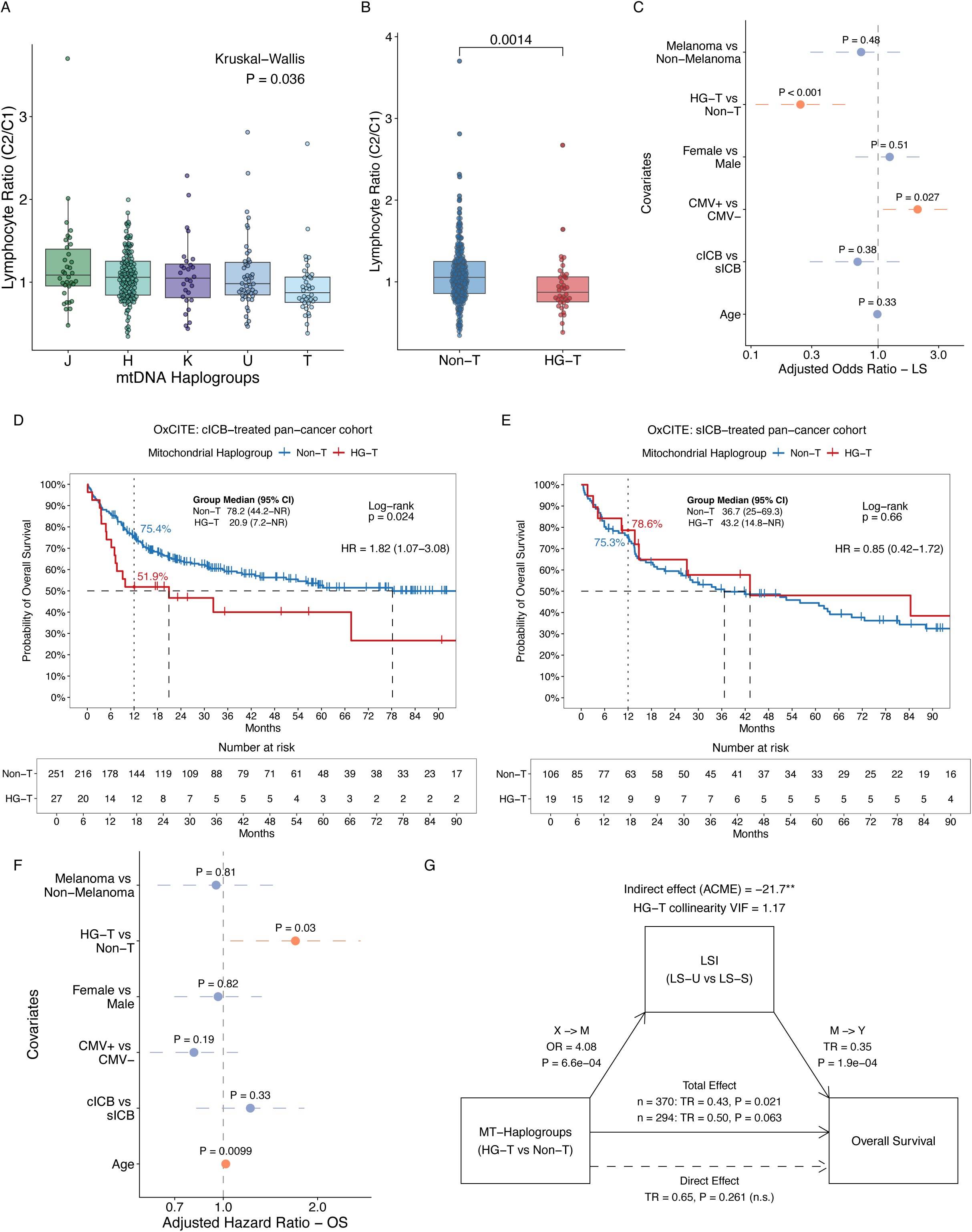
LSI mediates the effects of mitochondrial haplogroup T on ICB efficacy. **(A)** Lymphocyte ratio (C2/C1) across mitochondrial haplogroups J, H, T, U and K (Kruskal–Wallis test). **(B)** Lymphocyte ratio in haplogroup T (HG-T) versus Non-T patients in pan-cancer OxCITE cohort. **(C)** Multivariable logistic regression of LS adjusted for age, sex, cancer type, HG-T, CMV serostatus and regimen. **(D-E)** Kaplan– Meier curve showing OS by HG-T status in combination and single-agent ICB pan-cancer cohorts, respectively. **(F)** Multivariable Cox regression of OS adjusted for cancer type, HG-T, sex, CMV, regimen and age. **(G)** Mediation analysis evaluating LS as a potential mediator of the haplogroup T-OS association, showing path coefficients and corresponding P values for haplogroup T-LS association (X◊M) and LS-OS association (M◊Y), total/direct effects and average causal mediation effect (ACME). Variance inflation factors (VIF) indicated no substantial HG-T-associated collinearity.

#### HG-T predicts worse survival in the context of combination ICB

Given the association of HG-T carriage with LS, the relationship between HG-T carriage and OS was examined. Consistent with an association with increased risk of LS-U, HG-T carriage was likewise associated with poorer OS among all patients receiving cICB, whereas no significant association was observed in patients treated with single-agent ICB (sICB) (Figures 4D and 4E). Concordant observations were apparent when analysis was restricted analysis to patients with MM (Figures S5F and S5G). Multivariable Cox proportional hazards analysis further demonstrated that HG-T remained independently associated with worse OS (HR 1.70, 95% CI 1.05-2.74) after adjustment for age, sex, cancer type, treatment modality, and CMV serostatus (Figure 4F), supporting HG-T as an independent prognostic factor following cICB. These findings therefore suggest that the functional consequences of mitochondrial genetic background become more apparent under the heightened metabolic stress imposed by combination immunotherapy. Consistent with this interpretation, the association between HG-T and reduced lymphocyte ratio was likewise most evident in the cICB-treated cohort and was not observed in patients receiving sICB alone (Figures 4B and S5C).

#### LS partially mediates the inferior survival of HG-T carriers

Given that LS-U was associated with both HG-T carriage and shorter OS, we next asked whether the adverse clinical impact of HG-T is mediated through its effect on LS. To address this, we performed causal mediation analysis, modelling LS status by logistic regression, HG-T as the exposure and OS by a Weibull accelerated failure time (AFT) model. Both models were adjusted for age, sex, CMV serostatus, cancer type, and ICB regimen. In the full cohort, HG-T carriage was associated with shorter OS (time ratio [TR] 0.43, 95% CI 0.21-0.88, P = 0.021; n = 370). Among patients with available LS data (n = 294), in whom the mediation analysis was performed, the effect of HG-T on OS was of similar magnitude, albeit with borderline significance (TR 0.50, 95% CI 0.24-1.04, P = 0.063). Within this subset, HG-T carriers had approximately four-fold higher odds of LS-U (OR 4.08, 95% CI 1.80-9.22, P = 6.6×10^-4^). In turn, LS-U was independently associated with shorter OS (TR 0.35, 95% CI 0.20-0.61, P = 1.9×10^-4^). After LS status was included in the survival model, the association between HG-T and OS was attenuated and no longer significant (TR 0.65, 95% CI 0.31-1.37, P = 0.26).

Causal mediation analysis identified a significant indirect effect of HG-T on OS acting through LS. This effect corresponded to an average reduction of approximately 22 months in model-predicted survival time (average causal mediation effect [ACME] −21.7 months, 95% CI -47.0 to -5.5, P = 0.004), whereas the direct effect of HG-T was not significant (average direct effect [ADE] -38.2 months, 95% CI -97.7 to 39.1, P = 0.25) (Figure 4G). Consistent with this, the survival difference between HG-T and Non-T patients was attenuated after stratification by LS status, both among LS-U patients (HR 1.21, 95% CI 0.62-2.40, P = 0.57) and LS-S patients (HR 1.13, 95% CI 0.59-2.17, P = 0.72) (Figures S5H and S5I). Together, these results are consistent with LS-U forming an intermediate phenotype through which carriage of mitochondrial HG-T contributes, at least in part, to inferior outcomes following ICB.

### Divergent CD8+ T-cell transcriptional states between HG-T and Non-T

#### ICB Elicits Haplogroup-Specific Transcriptional Programs

To investigate whether mitochondrial haplogroup T influences transcriptional response to ICB, ICB-induced transcriptional changes in CD8+ T cells were separately characterised across HG-T and Non-T individuals. Similar to the transcriptional ICB response observed in patients with LS-U, HG-T carriers showed increased magnitude of ICB-responsive CD8+ T-cell transcriptional remodelling (Figure 5A, slope = 1.259, R² = 0.949; Table S7), reflecting a systematic amplification of both upregulated and downregulated DEGs directly due to HG-T carriage. Gene set enrichment analysis (GSEA) revealed that, whilst canonical T-cell activation pathways were enriched following ICB across all haplogroups, several programs were preferentially induced in HG-T carriers (Figure 5B and Table S8). These included mitochondrial electron transport chain activity (GO:0006123, 0006120), oxidative phosphorylation (GO:0006119), proton motive force-driven ATP synthesis (GO:0015986), and ATP metabolic process (GO:0046034), collectively indicating enhanced engagement of mitochondrial bioenergetic pathways by ICB in the CD8+ T cells for HG-T carriers. In parallel, pathways associated with intrinsic and receptor-mediated apoptosis (GO:2001233, 2001242, 1902041) were also upregulated in HG-T carriers, consistent with the central role of mitochondria in coordinating both cellular energy demands and stress-responsive cell death programs^32^.

**Figure 5:**
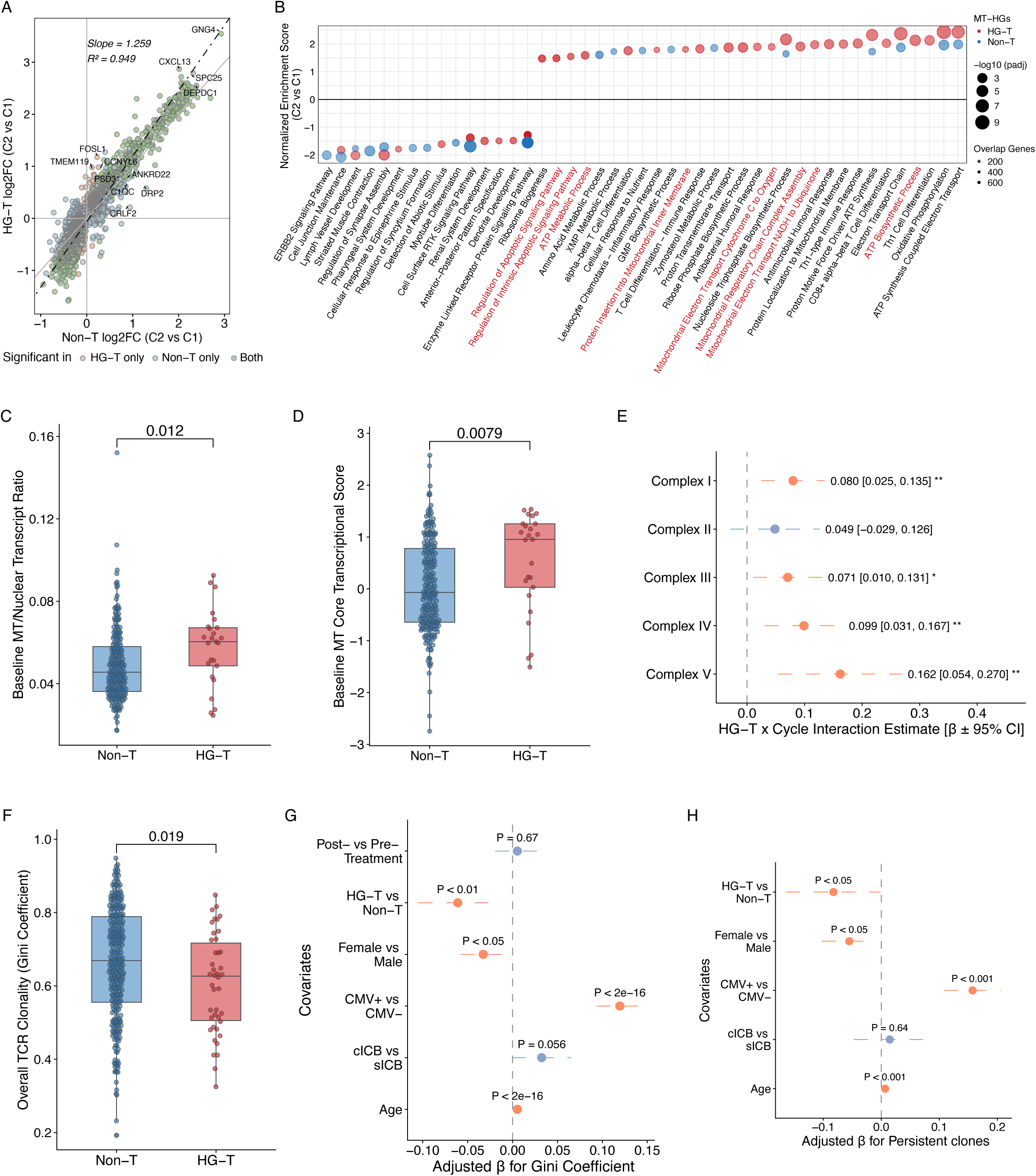
Haplogroup T-associated transcriptional and clonal features in response to ICB. **(A)** Pre- to post-ICB gene expression changes in HG-T and Non-T patients, indicating group-specific and shared significant genes. **(B)** GSEA of ICB-induced transcriptional changes in HG-T and non-T patients, ranked by between-group differences in normalized enrichment scores; dot size indicates significance. **(C)** Baseline mitochondrial-to-nuclear transcript ratio following variant-aware realignment. **(D)** Baseline core mitochondrial transcriptional Z-score, calculated as the sample-wise median of Z-standardized log2(TPM + 1) expression across the 13 mtDNA-encoded protein-coding genes. **(E)** HG-T × ICB interaction effects across respiratory chain complexes. **(F)** TCR clonality (Gini coefficient) by HG-T status. **(G-H)** Multivariable regression of TCR clonality and persistent CD8+ T-cell clone proportions, respectively.

#### HG-T CD8+ T exhibits a transcriptional hypermetabolic state

We next investigated whether HG-T carriage was associated with transcriptional differences in CD8+ T cells. To maximise statistical power while accounting for treatment status and population structure, HG-T and Non-T samples were compared across all available CD8+ T cell bulk RNA-seq profiles, correcting for treatment timepoints. Although genome-wide differences were modest at FDR<0.05, the strongest association was increased expression of *MTND2P28* in HG-T carriers (Figure S6A and Table S9). Analogous analyses of independently profiled B and NK cells identified the same association, albeit less significant, in other cell-types (Figure S6A), indicating the *MTND2P28* signal with HG-T carriage was reproducible across immune cell lineages, as would be expected if directly linked to haplogroup. Given that *MTND2P28* is a nuclear mitochondrial DNA segment (NUMT) homologous to mitochondrial *MT-ND2*, together with a reported susceptibility to read misalignment^33^, we reasoned that this signal might reflect haplogroup-dependent misassignment – reflecting genetic divergence from the reference sequence – rather than authentic pseudogene transcription. To test this a mitochondrial variant-aware reference genome was constructed, incorporating conservative haplogroup-associated mtDNA variations (allele frequency [AF] ≥ 0.01), and the RNA-seq data competitively re-aligned against the nuclear and mitochondrial genomes. Tracking fragments between the original and variant-aware alignments revealed substantial reassignment of reads originally mapping to *MTND2P28* to mitochondrial *MT-ND2*, while the majority of reads originally assigned to *MT-ND2* remained mitochondrial (Figure S6B). Thus, accounting for mitochondrial genetic variation substantially reduced systematic misassignment between mtDNA and homologous NUMTs.

To address whether resolving this mapping ambiguity uncovered a distinct HG-T associated transcriptional phenotype in patient-derived CD8+ T cells, the ratio of mitochondrial-to-nuclear transcripts was characterised. This demonstrated that, compared to other haplogroups, pre-treatment HG-T CD8+ T cells had an increased ratio of mitochondrial-to-nuclear transcripts (P = 0.012, Figure 5C). At the gene level, this difference extended beyond *MT-ND2,* with expression across the 13 mtDNA-encoded protein-coding genes, spanning respiratory complexes I, III, IV, and V, consistently increased in HG-T carriers (Figure S6C and Table S10). Reflecting this, a prespecified mitochondrial core transcriptional score summarising expression of the 13 mtDNA-encoded protein-coding genes was significantly elevated in HG-T CD8+ T cells at baseline (P = 7.9×10^-3^, Figure 5D). Together, these findings indicate that, in patients with cancer, HG-T CD8+ T cells enter ICB treatment with increased mitochondrial transcriptional activity. We further explored this divergent expression at mitochondrial complex-level analysis of nuclear-encoded genes (Table S11), reasoning that these should also track mitochondrial encoded changes. This demonstrated increased expression of mitochondrial programs across haplogroups following ICB (Figure S6D), but the magnitude of this induction for electron transport chain complexes I, III, and IV, together with ATP synthase (complex V), was significantly greater in HG-T individuals^34^ (Figure 5E). In summary, these results demonstrate that HG-T carriers – at least in the context of the immune exhaustion of cancer – demonstrate subtle but significant upregulation of the majority of mitochondrially encoded transcripts, reproducible across cell subsets, but most notable in CD8+ T cells. Moreover, ICB elicits induction of respiratory chain transcripts, but the effect of this is greater in HG-T carrying patients, consistent with an exaggerated mitochondrial transcriptional response to ICB in HG-T carriers.

#### HG-T Constrains Clonal Persistence and Reshapes the T Cell Repertoire

Given the transcriptional changes indicative of increased activation of apoptotic pathways in HG-T carriers, we examined the bulk CD8+ T-cell RNA-seq derived TCR repertoire data to assess the impact of HG-T carriage on T-cell clonotype dynamics. Using multivariable analysis, we observed that both age and CMV seropositivity were associated with higher Gini coefficients, reflecting the known effect of ageing as well as CMV related memory inflation^35^. Conversely, biological female sex and, most significantly, carriage of HG-T was associated with lower Gini coefficients, in keeping with reduced repertoire clonality (Figures 5F and 5G). Intriguingly, the effect of HG-T carriage remained significant after accounting for LS (Fig S6E, LS P = 0.27, HG-T P <0.05), indicating that mitochondrial HG-T carriage directly accounted for divergence in TCR clonality observed in patients, irrespective of LS. To assess whether the apoptotic signaling observed in HG-T carriers had discernible effects on T-cell clonal survival, longitudinal samples collected at each cycle of ICB were examined. This showed that HG-T carriers exhibited significantly fewer persistent clones after one cycle of ICB, with a corresponding increase in clones lost following treatment compared to Non-T carriers (Figures 5H and S6F). Thus, the enhanced apoptotic signaling observed in the HG-T transcriptional landscape translated to detectable effects on the TCR repertoire, characterised by reduced representation of expanded memory-like clones and reduced clonal stability, in keeping with impaired CD8+ T cell clonal stability in HG-T carriers.

### HG-T-specific transcriptional and clonal features at single-cell resolution

#### HG-T CD8+ T Cells Exhibit a Selective Loss of TPEX States

To provide granularity to these observations and further characterise the CD8+ T-cell subsets showing most transcriptional sensitivity to HG-T carriage, a peripheral CD8+ T-cell atlas comprising 5’ scRNA-seq with paired VDJ sequences was generated. Post-quality control, this comprised 220,060 cells across 195 pre- and post-treatment samples from 120 patients receiving ICB for melanoma (Figure 6A). CD8+ T cells were annotated into distinct functional subsets, including naïve (CD8.Naive), central memory (CD8.TCM), effector memory (CD8.TEM), terminally differentiated effector memory (CD8.TEMRA), stem-like progenitor exhausted (CD8.TPEX), proliferative (CD8.Pro), mucosal-associated invariant T (MAIT), gamma-delta T (gdT), and mitochondrial gene expression-high (CD8.Mitohi) populations (Figure 5A). Canonical marker expression and quality control metrics were consistent with established subset identities (Figures S7A – S7E). Comparative analysis of subset proportions between HG-T carriers versus non-carrier patients using a linear mixed-effects model, adjusting for treatment timepoints and inter-individual variance, showed that all subsets showed similar proportions across haplogroups except for CD8.TPEX cells, which were found at a reduced frequency in HG-T carriers (P = 0.03; Figure 6B). A similar result was obtained through per-sample Wilcoxon tests at baseline, with a consistent direction of effect noted post-treatment (C1, P = 0.029; C2, P = 0.066; Figure S8A and Table S12). To test whether this observation was driven by outlier individuals, a leave-one-donor-out sensitivity analysis was performed across the 12 HG-T individuals. For all iterations the odds ratio remained below one however, with 7 out of 12 iterations reaching statistical significance with this smaller sample and the remainder showing borderline significance (Figure S8B), demonstrating fewer CD8.TPEX cells across HG-T carriers irrespective of donor heterogeneity. It is increasingly clear that such stem-like TPEX cells are crucial for sustaining CD8+ T-cell responses upon ICB, and these results indicate HG-T carriers have a compromised reservoir for T-cell reinvigoration, mirroring the impaired LS in these patients^36,37^. Finally, the molecular basis underlying reduced TPEX proportions was explored with single-cell regulatory network inference using pySCENIC. This identified five CD8.TPEX-specific regulons, with the *MYB(+)* regulon emerging as the most specific to this subset (Figure S8C), in keeping with the described crucial role of the MYB transcription factor in the regulation of self-renewal and maintenance of stem-like CD8+ T cells^38,39^. Thus, the association of HG-T carriage with reduced CD8+ T cell clonal maintenance was supported by specific reductions in the proportion of CD8.TPEX cells, characterised by expression of persistence-related regulatory networks and central to durable ICB responses (Figure S8D).

**Figure 6:**
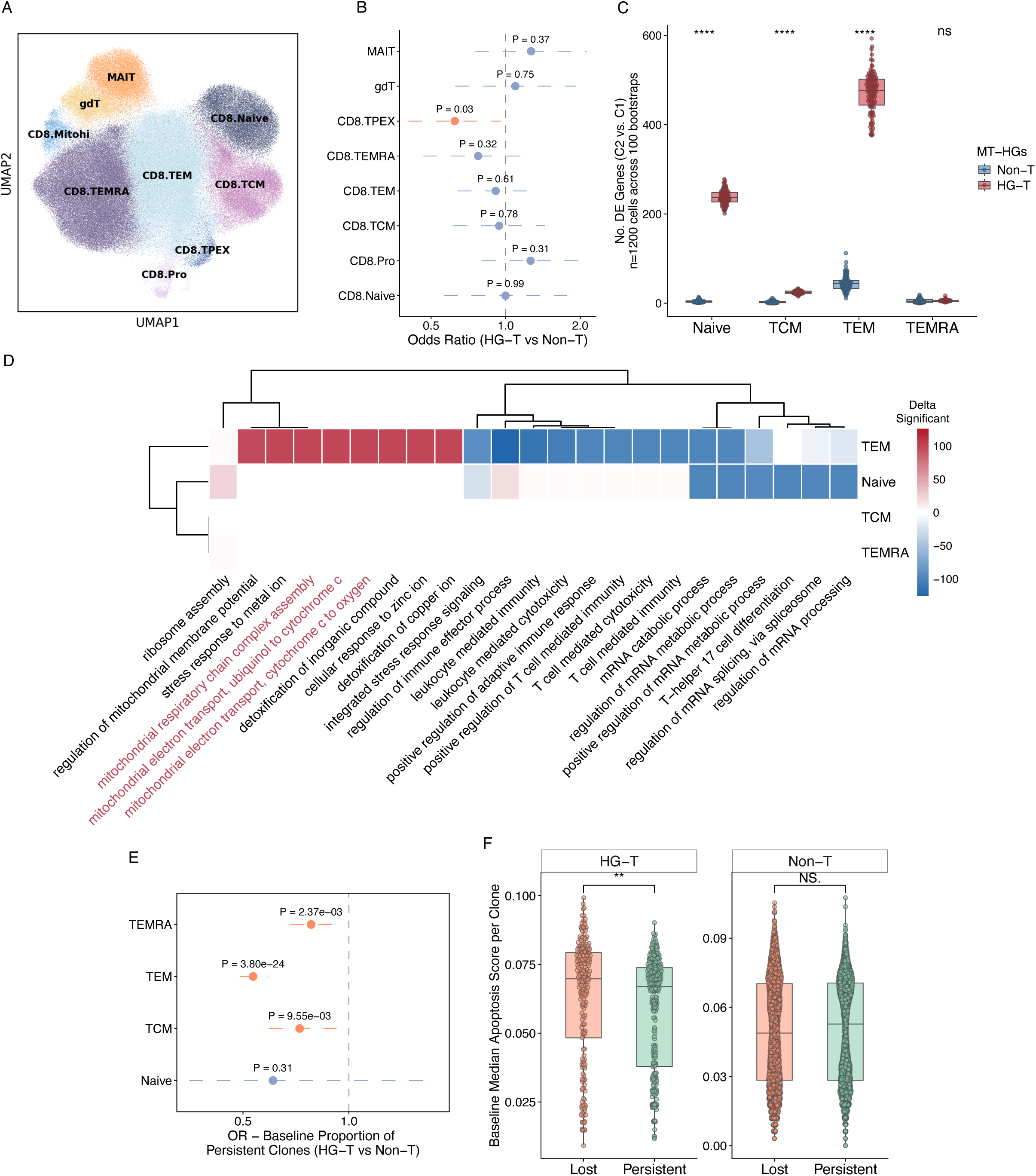
Single-cell characterization of haplogroup T-associated immune landscape. **(A)** UMAP visualization of CD8+ T-cell subsets (naïve, CM, EM, TEMRA, MAIT and γδ). **(B)** Association of HG-T with CD8+ T-cell subset abundance (OR, 95% CI). **(C)** Number of ICB-induced differentially expressed genes (DEGs) across functional CD8+ T-cell types and HG-T status (bootstrap-based Wilcoxon tests). **(D)** GOBP enrichment of longitudinal changes before and after ICB, coloured by between-group differences in significant bootstrap iterations. **(E)** Association of HG-T with baseline persistent clone proportions across CD8+ T-cell states (OR, 95% CI). **(F)** Baseline apoptosis scores in lost versus persistent clones by HG-T status.

#### TEM Links Transcriptional Rewiring to Clonal Dynamics in HG-T

The bulk CD8+ T cell RNA-seq analysis indicated that, in keeping with effects of LS-U, HG-T carriage was associated with exaggerated transcriptional response to ICB. To determine whether this divergence was reflected across distinct CD8+ T-cell states, ICB-induced DE analysis was examined across naive, TCM, TEM and TEMRA subsets. To ensure balanced comparisons, equal numbers of cells from HG-T and Non-T carrying patients were subsampled within each subset and bootstrap-based DE analyses performed (C2 vs C1). This permitted non-parametric identification of DEGs while controlling for differences in statistical power across subsets. We complemented this approach with independent validation using donor-balanced, linear mixed-effects models. Across all CD8+ T-cell subsets, HG-T carriers consistently exhibited exaggerated transcriptional responses to ICB, with naïve and TEM subsets showing the greatest divergence from Non-T individuals, as reflected by the largest differences in the number of DEGs (Figures 6C and S9A, Table S13). Notably, naïve cells were characterized predominantly by transcriptional downregulation, consistent with disruption of baseline homeostatic programs, whereas TEM cells showed haplogroup T-specific upregulation of mitochondrial metabolic pathways (Figure 6D and Table S14). Examination of canonical markers further supported these observations with *BCL2*, a key anti-apoptotic factor, being preferentially downregulated in HG-T naïve cells, while *HLA-DRA* and *IL7R* exhibited increased and decreased expression, respectively, in HG-T TEM cells (Figure S9B), consistent with enhanced activation coupled with reduced memory potential.

We next revisited clonal persistence across subsets, defining clones through paired alpha-beta TCR sequences derived from single-cell V(D)J sequencing. In keeping with observations from bulk RNA-seq data, cells derived from persistent clones were significantly reduced in HG-T patients across TCM, TEM and TEMRA populations, with the strongest effect observed in TEM (Figure 6E). The robustness of these observations to outlier individuals was further assessed using repeated subsampling of paired samples (8 HG-T and 8 Non-T donors) across 1,000 bootstrap iterations. At the cell level, the reduction in persistent clones remained significant in ∼82.4% of iterations for TEM, whereas less than 50% of iterations reached significance in TCM or TEMRA (Figure S9C), suggesting that the effect is most consistently driven by the reduced clonal persistence in the TEM compartment.

To determine whether clonal fate was influenced by transcriptional state, we compared baseline transcriptional profiles of clones that were resampled, consistent with clonal persistence, versus those that were not at C2. In Non-HG-T carriers, metabolic and apoptotic features did not distinguish between persistent and non-persistent clones, indicating other pressures for clonal persistence, most likely antigen recognition. In contrast, HG-T clones that were lost to sampling post ICB exhibited markedly elevated expression of apoptosis, ROS, glycolysis, and hypoxia signatures pre-treatment (Figures 6F and S9D). These findings indicate that CD8+ T cells from HG-T carriers demonstrate proportionally increased metabolic stress prior to treatment, resultant in shorter clonal half-life.

## Discussion

Many established correlates of response to ICB – including the maintenance of large peripheral CD8+ T-cell clones over treatment and an intact stem-like progenitor compartment crucial for sustaining effector activity^14,40^ – reflect properties of the host T-cell compartment. Treatment selection in MM however is based on clinical and tumour-intrinsic factors, and does not take account host T-cell fitness. Clinically, pre-emptive identification of ICB non-responders is attractive for its potential to identify non-responding patients early so as to enable treatment switching and escalation. Here we show that LS – lymphocyte stability – representing the ratio of lymphocytes derived from routine blood tests across the first cycle of ICB, identifies patients with impaired long-term survival in MM well in advance of standard radiological assessment timepoints. We further show that LS reflects underlying divergent CD8+ T-cell mitochondrial activity, with an exaggerated oxidative transcriptional response to treatment characterising poorer outcomes. Notably, LS is in part determined by inherited mitochondrial variation: carriage of mitochondrial haplogroup T (HG-T) being associated with ICB hyperresponsiveness, reduced progenitor-exhausted TPEX compartment, impaired T-cell clonal persistence and shortened OS.

Lymphocyte stability (LS), the preservation of peripheral lymphocyte counts after one cycle of ICB, is a dynamic on-treatment indicator of OS across anti-PD-1 and anti-CTLA-4 regimens. After adjustment for clinical covariates, LS remains prognostic and further stratifies patients with elevated baseline NLR, suggesting that it captures on-treatment prognostic information distinct from baseline indices. Paired CD8+ T cell RNA-seq pre- and post-ICB revealed a substantially greater response in LS-U patients (more DEGs, larger effects), preferentially involving oxidative phosphorylation and electron transport chain pathways. Conversely, LS-S-associated pathways were characteristically anabolic, including positive regulation of mitochondrial translation and IL-7 signalling, consistent with preserved biosynthesis and homeostatic survival. In addition, although PD-1 signalling regulates T-cell glycolysis, fatty-acid oxidation and oxidative activity^44^, the bioenergetic effects of checkpoint blockade in patient T cells remain incompletely characterised^45^. In primary patient samples, ICB consistently increased OCR, ATP production, as well as maximal and spare respiratory capacity. These data provide direct evidence that ICB is accompanied by invigorated mitochondrial activity, raising the possibility that inter-individual differences in adapting to energetic demand contribute to LS.

Given the association between LS and mitochondrial pathways, we reasoned that germline mitochondrial variation might influence LS. Mitochondrial haplogroups comprise hundreds of linked, co-evolved variants that influence mitochondrial biology^20^. Testing haplogroup against LS revealed a strong association between HG-T carriage, present in ∼10% of the white UK population^20^, and LS-U phenotype. Consistent with the recently reported impaired radiological response to ICB among HG-T carriers with MM^27^, we find that HG-T carriage was associated with reduced OS in our independent, prospective OxCITE cohort. This association was at least partly mediated through LS, with no haplogroup effect on survival among LS-matched patients, positioning LS as an intermediate phenotype linking inherited mitochondrial variation with treatment outcome. These findings support HG-T as an upstream contributor to lymphocyte instability, but not the only determinant of LS-U.

Initial analyses identified *MTND2P28* as the strongest HG-T-associated transcript across CD8+ T, B and NK cells (adjusted P = 7.1×10⁻²⁰, 4.0×10⁻⁵ and 1.6×10⁻¹⁹, respectively). However, *MTND2P28* is a nuclear mitochondrial DNA segment (NUMT) homologous to mitochondrial *MT-ND2*, and we observed implausibly high read counts for a pseudogene when using a single reference mitochondrial genome. Because NUMTs create alignment ambiguity, particularly when inherited mtDNA variation increases divergence from a single mitochondrial reference sequence^46,47^, we therefore realigned reads to a variant-aware reference. This reassigned many pseudogene-mapped reads to the mitochondrial genome across all haplogroups, demonstrating that inherited mtDNA variation can generate genotype-dependent alignment artefacts. After remapping, HG-T carriers showed coordinated baseline elevation of mtDNA-encoded protein-coding transcripts in CD8+ T cells. Because germline effects attenuate with molecular distance from the causal variant^48,49^, a causal haplogroup effect should be most detectable in genes governing mitochondrial function, with immediate secondary effects on metabolic pathways. Consistent with this, beyond mtNDA-encoded transcripts, HG-T carriers showed differential expression of the nuclear-encoded metabolic genes *SLC16A14* and *NXN*, supporting extension of the HG-T phenotype to metabolic and redox pathways through mitochondrial-nuclear crosstalk^50^. These baseline effects were nevertheless relatively modest, suggesting a subtle genotype influence that manifests under the immunological demands of metastatic cancer and ICB. Indeed, ICB induced nuclear-encoded oxidative phosphorylation programmes across haplogroups, but more strongly upregulated in HG-T across respiratory-chain complexes I, III, IV and V.

Failure of homeostatic maintenance of lymphocyte populations after ICB may reflect increased cell death or impaired progenitor-derived replenishment. A single-cell RNA and VDJ atlas of >220,000 CD8+ T cells from 120 patients (75 paired pre- and post-ICB) revealed three further key attributes of HG-T carriage that were consistent with bulk RNA-seq observations. Primarily, HG-T was associated with an exaggerated response to ICB, particularly in CD8+ TEM cells, with over-representation of induction of genes implicated in mitochondrial electron transport. Secondly, while we did not replicate the expanded naïve T-cell populations reported in a previous pre-treatment dataset where analyses were split according to clinical benefit defined by radiological response^27^, we did see an association between HG-T carriage and pre-treatment CD8.TPEX proportion, with a similar post-ICB trend. Enrichment of a *MYB*-driven regulon in these cells, given the critical role of MYB in anti-PD-1-triggered TPEX proliferative bursts^38^, strongly supports impaired homeostatic proliferation. Finally, ICB specifically upregulated apoptosis-related pathways in HG-T CD8+ T cells, consistent with mitochondrial control of apoptosis^51^. Homeostatic T-cell turnover is a response-associated feature of ICB^52^; sc-VDJ tracking showed baseline clones in HG-T carriers were less likely to be re-detected after treatment, and non-persistent clones showed baseline enrichment of apoptosis and ROS pathways.

We propose a priming-activation-attrition model to integrate these observations. For HG-T, inherited mitochondrial variation establishes a primed baseline state, upon which ICB imposes increased bioenergetic demand; the greater induction of respiratory-chain programmes in HG-T cells may therefore reflect increased demand or compensatory adaptation rather than greater mitochondrial fitness. Baseline CD8.TPEX depletion with reduced MYB-associated programmes is consistent with this state being one of increased cellular stress. After ICB, increased apoptotic programmes and reduced clonotype persistence suggest that altered metabolic adaptation in HG-T favours activation and differentiation at the expense of progenitor maintenance and clonal persistence.

Our study thus identifies LS as an integrative on-treatment phenotype, linking clinical outcome with T-cell metabolism and clonal persistence during ICB, derivable from routine blood testing and therefore potentially scalable for dynamic monitoring during therapy. The association of HG-T with lymphocyte instability provides evidence that inherited mitochondrial variation can shape T-cell capacity to meet the metabolic demands of ICB. Metabolic adaptability, rather than activation alone, is a potential determinant of sustained immunity - which may help explain why strong effector activation does not necessarily translate into durable response. As inhibition of additional checkpoints has yielded limited improvement beyond PD-1-based therapy^53^, our findings build on recent work that suggests approaches to support T-cell mitochondrial function^54^, rather than further releasing inhibitory brakes, may represent a complementary strategy to enhance ICB.

### Limitations of the study

First, the study remains predominantly observational. HG-T is present in only a subset of LS-U patients, indicating that it represents one inherited contributor rather than a deterministic cause of lymphocyte instability. Other host-, tumour-, and treatment-related factors likely contribute to LS. Second, although our transcriptional and cellular findings are consistent with altered mitochondrial adaptation in HG-T CD8+ T cells, they do not directly establish mitochondrial dysfunction. Seahorse experiments assessed *ex vivo* stimulated T cells, rather than tumour-reactive CD8+ T cells, and were underpowered for HG-T-specific phenotypes. Further functional comparisons are required to distinguish compensatory adaptation and intrinsic respiratory dysfunction. Finally, short-read sequencing cannot completely resolve homologous mtDNA and NUMT sequences. Although our variant-aware alignment substantially reduced genotype-dependent read misassignment, residual ambiguous mapping is likely to remain. Long-read sequencing, ideally incorporating haplotype-resolved mitochondrial and nuclear genomes, will be important for validating haplogroup-associated mitochondrial transcription.

## STAR ★ Methods

### KEY RESOURCES TABLE

| REAGENT or RESOURCE | SOURCE | IDENTIFIER |
|---|---|---|
| Biological samples |  |  |
| PBMCs, human (OxCITE) | Oxford University Hospitals NHS Foundation Trust | See Table S1 |
| PBMCs, human (External Validation) | Oxford University Hospitals NHS Foundation Trust | See Table S2 |
| Antibodies |  |  |
| Ultra-LEAF™ Purified anti-human CD3 Antibody (Clone: UCHT1) | BioLegend | 300438 |
| Ultra-LEAF™ Purified anti-human CD28 Antibody (Clone: CD28.2) | BioLegend | 302934 |
| Pan T cell Isolation Kit, human | Miltenyi Biotec | 130-096-535 |
| CD8 Microbeads, human | Miltenyi Biotec | 130-045-201 |
| CD19 Microbeads, human | Miltenyi Biotec | 130-050-301 |
| Chemicals, peptides, and recombinant proteins |  |  |
| Poly-D-Lysine 0.1mg/ml | Gibco | A38904-01 |
| Critical commercial assays |  |  |
| Seahorse XF T Cell Metabolic Profiling Kit | Agilent | 103772-100 |
| Seahorse XF RPMI Medium, pH 7.4 | Agilent | 103576-100 |
| Seahorse XF 1.0 M Glucose Solution | Agilent | 103577-100 |
| Seahorse XF 100 mM Pyruvate Solution | Agilent | 103578-100 |
| Seahorse XF 200 mM Glutamine Solution | Agilent | 103579-100 |
| Software and algorithms |  |  |
| bamtools v2.5.1 | Barnett et al.<sup>55</sup> | https://github.com/pezmaster31/bamtools |
| Cell Ranger v8.0.1 | 10X Genomics | https://www.10xgenomics.com/support/software/cell-ranger/latest |
| Cellsnp-lite v1.2.0 | Huang et al.<sup>56</sup> | https://cellsnp-lite.readthedocs.io/en/latest/ |
| CellTypist v1.7.1 | Domínguez Conde et al.<sup>57</sup> | https://www.celltypist.org/ |
| clusterprofiler v4.18.4 | Xu et al.<sup>58</sup> | https://github.com/YuLab-SMU/clusterProfiler |
| DESeq2 v1.50.2 | Love et al.<sup>59</sup> | https://bioconductor.org/packages/release/bioc/html/DESeq2.html |
| DropletUtils v1.28.0 | Lun et al.<sup>60</sup> | https://github.com/MarioniLab/DropletUtils |
| fgsea v3.22 | Korotkevich et al.<sup>61</sup> | https://github.com/alserglab/fgsea |
| GenomeStudio v2.0.5 | Illumina | https://emea.support.illumina.com/array/array_software/genomestudio/downloads.html |
| Haplogrep3 v3.2.1 | Schonherr et al.<sup>62</sup> | https://haplogrep.i-med.ac.at/ |
| HISAT2 v2.2.1 | Kim et al.<sup>63</sup> | https://github.com/DaehwanKimLab/hisat2 |
| HTSeq v2.0.9 | Anders et al. <sup>64</sup> | https://github.com/htseq/htseq |
| IMPUTE2 v2.3.2 | Howie et al.<sup>65</sup> | https://mathgen.stats.ox.ac.uk/impute/impute_v2.html |
| lme4 v1.1-38 | Bates et al.<sup>66</sup> | https://github.com/lme4/lme4 |
| mediation v4.5.1 | Tingley et al.<sup>67</sup> | https://cran.r-project.org/web/packages/mediation/index.html |
| MiXCR v3.0.13 | Bolotin et al.<sup>68</sup> | https://mixcr.com/ |
| Picard v3.1.1 | Broad Institute | https://broadinstitute.github.io/picard/ |
| Plink v1.9b6.24 | Chang et al.<sup>69</sup> | https://www.cog-genomics.org/plink/ |
| pROC v1.19.0.1 | Robin et al.<sup>70</sup> | https://cran.r-project.org/web/packages/pROC/index.html |
| pySCENIC v0.12.2b0 | Aibar et al.<sup>71</sup> | https://github.com/aertslab/SCENIC |
| samtools v1.17 | Li et al.<sup>72</sup> | https://github.com/samtools/samtools |
| Scanpy v1.11.5 | Wolf et al.<sup>73</sup> | https://scanpy.readthedocs.io/en/stable/ |
| scDblFinder v1.22.0 | Germain et al.<sup>74</sup> | https://github.com/plger/scDblFinder |
| scIB-metrics v0.5.7 | Gayoso et al.<sup>75,76</sup> | https://scib-metrics.readthedocs.io/en/stable/ |
| scVI-tools v1.1.5 | Lopez et al.<sup>77</sup> | https://scvi-tools.org/ |
| Seurat v5.4.0 | Hao et al.<sup>78</sup> | https://satijalab.org/seurat/ |
| soupX v1.6.2 | Young et al.<sup>79</sup> | https://github.com/constantAmateur/SoupX |
| survival v3.8-3 | CRAN | https://cran.r-project.org/web/packages/survival/index.html |
| survminer v0.5.1 | CRAN | https://cran.r-project.org/web/packages/survminer/index.html |
| UCell v2.14.0 | Andreatta and Carmona<sup>80</sup> | https://github.com/carmonalab/Ucell |
| Vireo v0.5.8 | Huang et al.<sup>81</sup> | https://vireosnp.readthedocs.io/en/latest/ |

## EXPERIMENTAL MODEL AND SUBJECT DETAILS

### Oxford Cancer Immunotherapy Toxicity and Efficacy (OxCITE) Cohort

This study involves human participants and was approved by Oxford Radcliffe Biobank (Oxford Centre for Histopathology Research ethical approval reference 19/SC/0173, project nos. 16/A019, 18/A064 and 19/A114). Participants gave informed consent to participate in the study before taking part.

Patients eligible for immune checkpoint blockade, including both palliative and adjuvant settings, at Oxford University Hospitals NHS Foundation Trust were prospectively recruited between 23 November 2015 and 25 July 2025. The OxCITE cohort comprised 417 patients, of whom 406 has advanced or metastatic cancer, including 353 with melanoma, 24 with renal cell carcinoma, 15 with colorectal cancer, and 25 with mesothelioma. Patients received either combination ICB (cICB) (n = 279; ipilimumab plus nivolumab) or single-agent ICB (sICB) (n = 127; pembrolizumab, n = 109; nivolumab, n = 18). Immunological and clinical outcome analyses were restricted to the 406 advanced or metastatic patients treated with either sICB or cICB across cancer types. To minimise confounding factors due to tumour-type heterogeneity, all bulk and single-cell transcriptomic analyses were further restricted to melanoma patients receiving sICB or cICB, as detailed below. An additional 11 melanoma patients receiving adjuvant pembrolizumab (adjICB) were incorporated in Seahorse metabolic profiling analysis to increase statistical power for haplogroup-based comparisons. Cohorts included for each figure are listed in Tables S1 and S15.

### External validation cohort

For the validation cohort, the following data were retrospectively extracted from healthcare records for every patient treated with immune checkpoint blockade (systemic anticancer regimens of monoclonal antibodies acting on PD-1, PD-L1 or CTLA-4) at Oxford University Hospitals NHS Foundation Trust between 01-Jan-2014 and 20-Oct-2022 : sex (as defined in the medical record), age at treatment start, date of death (if no longer alive), treatment, treatment start date, treatment intent, treatment regimen administered, ICD-10 diagnosis, lymphocyte count (30 days prior and up to 50 days after treatment start date). This information was used to select cases where melanoma was treated palliatively with immune checkpoint blockade (i.e. excluding adjuvant intent treatment) and to calculate LSI and overall survival and perform downstream survival analysis as outlined below. Permission to obtain and use anonymised retrospective healthcare data without participant consent in this study was granted by the Health Research Authority and Health and Care Research Wales and sponsored by the University of Oxford Research Governance, Ethics and Assurance (IRAS reference 317078; REC reference 22/HRA/3763). Patient details are listed in Table S2.

## METHOD DETAILS

### Sample collection

Peripheral blood samples (30-50 mL) were collected prior to initiation of immune checkpoint blockade (ICB) and longitudinally before each subsequent treatment cycle. Plasma and peripheral blood mononuclear cells (PBMCs) were isolated by Ficoll-Paque density gradient centrifugation. PBMCs were processed as follows: (i) cells were lysed in RLT Plus buffer (Qiagen) supplemented with dithiothreitol (DTT; Merck) and snap-frozen at −80 °C for RNA and DNA extraction; (ii) immune cell subsets were enriched using magnetic-activated cell sorting (MACS) (Miltenyi Biotec) for downstream bulk and single-cell RNA sequencing; and (iii) aliquots were cryopreserved in 90% fetal calf serum (FCS; Gibco) with 10% dimethyl sulfoxide (DMSO; Merck) for future use. Peripheral blood plasma collected at baseline (day 0) was diluted 1:2 in HBSS and used to quantify CMV IgG titres on an Abbott Architect i2000 immunoassay platform. For all analyses, the blood sample obtained closest to treatment initiation (within -30 to 0 days) was used as the baseline time point.

#### Clinical outcomes and lymphocyte stability (LS) calculations

Patient demographic and clinical characteristics were extracted from the electronic patient record (EPR). Disease stage was assigned according to American Joint Committee on Cancer (AJCC) 8th edition staging system, with Tumour Node Metastasis (TNM) 8th edition criteria for metastasis subclassification (M1a-d). All patients included had a minimum follow-up duration of 6 months for survival analyses. PFS was defined as time of ICB initiation to clinical or radiological evidence of progression (according to irRECIST.1.1 criteria), or death. OS was defined as time from ICB initiation to death from any cause. Median follow-up duration was determined using a reverse Kaplan-Meier method.

To calculate lymphocyte stability (LS) and neutrophil-to-lymphocyte ratio (NLR), full blood counts were obtained from EPR, where they were generated via Sysmex XN series analyzers (Sysmex Corporation) in an NHS clinical diagnostic laboratory. Pre-treatment (C1) samples were defined as the most recent sample prior to ICB initiation (from Day -30 to Day 0), and post-treatment (C2) samples were the most recent sample after the first treatment cycle (from Day 18 to Day 50). Patients with a prior diagnosis of lymphoid-lineage haematological malignancies (e.g. chronic lymphocytic leukemia, lymphoma) were excluded.

Lymphocyte stability (LS) is defined as:

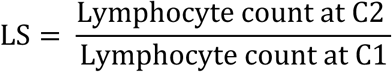

where LS-stable (LS-S) is defined as LS > 0.8, and LS-Unstable (LS-U) is defined as LS ≤ 0.8.

The optimal cutpoint for LS at 0.8 was determined by maximally selected log-rank statistics using *surv_cutpoint()* function from survminer (version 0.5.1). This was supported by time-dependent receiver operating characteristic (ROC) analysis using pROC (version 1.19.0.1), where the 24-month optimal cutoff was 0.804 based on maximal Youden index.

### Bulk RNA-seq preprocessing

Poly(A)-selected RNA libraries were sequenced using either 150-bp paired-end reads on an Illumina NovaSeq platform or 75-bp paired-end reads on an Illumina HiSeq 4000, both performed at the Oxford Genome Centre. Sequencing reads were aligned to the GRCh38/hg38 reference genome using HISAT2 (version 2.2.1), and gene-level read counts were generated with HTSeq (version 2.0.9). High-quality alignments were retained based on mapping quality (MAPQ) scores using bamtools (version 2.5.1). Duplicate reads were marked and removed using Picard (version 3.1.1), and samtools (version 1.17) was used for processing aligned reads and generating alignment statistics. A total of 246 baseline cycle 1 (C1) CD8⁺ T-cell transcriptomes (25 HG-T and 221 Non-T), 195 post-treatment cycle 2 (C2) transcriptomes (22 HG-T and 173 Non-T), and 67 post-treatment cycle 4 (C4) transcriptomes (5 HG-T and 62 Non-T) were included in the analysis. Among these, 174 donors (21 HG-T and 153 Non-T) were longitudinally paired across time points. Gene expression counts were normalised using DESeq2 (version 1.50.2).

### Competitive re-alignment with variant-aware reference genome

To account for sequence-dependent misalignment between mitochondrial transcripts and nuclear mitochondrial DNA segments (NUMTs), we generated a mitochondrial variant-aware alignment reference based on GRCh38. Mitochondrial variants associated with the haplogroups represented in the cohort, together with conservative mitochondrial variants (AF ≥ 0.01) catalogued in the Genome Aggregation Database (gnomAD) v3.1, were incorporated into the HISAT2 reference as SNP and haplotype information, while retaining the complete nuclear genome to permit competitive alignment between mitochondrial and homologous nuclear loci. HISAT2 indices were generated using hisat2-build with the --snp and --haplotype options, and paired-end RNA-seq reads were subsequently realigned to the resulting variant-aware reference using HISAT2. Gene-level expression was re-quantified from the variant-aware alignments using HTSeq-count with the same annotation and counting parameters applied to the original analysis. These re-derived counts were used for all downstream analyses involving mitochondrial and nuclear mitochondrial pseudogene expression. TPM values were calculated from the re-quantified gene counts using gene lengths derived from the corresponding GRCh38 annotation.

### Differential expression analysis

Differential expression (DE) analysis was performed using DESeq2 (version 1.50.2). Genes with low expression were filtered prior to analysis, retaining only those with a mean count > 10 across samples. For longitudinal comparisons between baseline (C1) and post-treatment cycle 2 (C2) samples, DE analysis was conducted in a paired manner using the subset of 174 matched samples, incorporating patient identity as a blocking factor in the design formula to account for inter-individual variability. For cross-sectional comparisons between HG-T and Non-T groups at each time point (baseline and post-treatment), the top three principal components derived from variance-stabilised expression data were included as covariates in the design matrix, unless otherwise specified, to mitigate potential technical variation.

We performed GSEA using fgsea (version 3.22) with MSigDB Gene Ontology Biological Process (GOBP) datasets. Genes were ranked by average log₂ fold change (FC) derived from DE analysis. To ensure comparability between contrasts, only overlapping genes between the two groups were retained as the background set. To minimise selection bias and enhance robustness, enriched pathways were ranked based on the difference in normalised enrichment scores (ΔNES) between groups, and the top-ranked pathways were selected for visualisation. Over-representation analysis was conducted using clusterprofiler (version 4.18.4) with MSigDB GOBP gene sets with DEGs from each comparison tested against the shared gene universe using a hypergeometric test with Benjamini-Hochberg correction for multiple testing. Enriched pathways were ranked by fold enrichment, and the highest ranking terms were visualised.

### Mitochondrial complex transcriptional activity

Mitochondrial complex-specific gene sets were curated from MSigDB (C2:CP:WikiPathways; complexes I-IV) and the HUGO Gene Nomenclature Committee (HGNC; complex V). Normalised gene expression values were extracted from baseline and post-treatment samples, and pathway activity scores were calculated as the geometric mean of gene expression within each complex. To assess the interaction between mitochondrial haplogroup status and treatment over time, linear mixed-effects models were fitted using patient ID as a random effect to account for repeated measurements. Fixed effects included mitochondrial haplogroup, treatment time point, and their interaction term.

### Bulk adaptive receptor analysis

T cell receptor (TCR) repertoire profiling was performed on bulk CD8+ RNA-seq data from paired pre- and post-treatment samples using MiXCR (version 3.0.13). Sequencing reads were aligned to reference V, D, and J gene segments to reconstruct TCR clonotypes based on complementarity-determining region 3 (CDR3) sequences. To improve clonotype assembly, the non-default parameter *OallowPartialAlignments = true* was enabled to retain partially aligned reads for downstream assembly. Three iterative rounds of partial read assembly (*assemblePartial*) were conducted to maximise recovery of CDR3-containing sequences. In addition, quality-aware error correction during clonotype assembly and clustering was applied using a threshold of *ObadQualityThreshold = 15*.

Samples with fewer than 500 detected TCR clones were excluded to ensure sufficient repertoire complexity, and one outlier C2 sample from patient ID 250 was removed from downstream analysis. Following quality control, a total of 431,709 TRA chains and 714,627 TRB chains were identified across 173 donors with paired samples, including 106 donors with C1/C2 pairs and 67 donors with C1/C2/C4 longitudinal sampling. Clonotypes were defined based on the amino acid sequence of the CDR3 region in combination with TRB V and J gene usage. Clone size was quantified using TRB chains, with clonal expansion measured as the proportion of total repertoire occupancy (clone read count / total TRB read count). Persistent clones were defined as clonotypes present in both pre-treatment (C1) and post-treatment (C2) samples from the same individual. Lost clones were defined as clonotypes present at C1 but absent at C2, whereas new clones were defined as clonotypes absent at C1 but detected at C2.

### Single-cell RNA-seq preprocessing, integration and annotation

Positively selected CD8⁺ T cells, together with peripheral blood mononuclear cells (PBMCs), were quantified and partitioned into nanoliter-scale droplets using the 10X Genomics Chromium platform, followed by cell lysis and reverse transcription. Complementary DNA (cDNA) was used to generate 5′ gene expression (GEX) and V(D)J libraries using the Chromium Next GEM Single Cell 5′ Reagent Kit v2 (Dual Index). Droplet encapsulation and reverse transcription were performed on individual samples, whereas library preparation and sequencing were conducted in batches. Sequencing was performed on an Illumina HiSeq 4000 platform, generating 75-bp paired-end reads for 5′ gene expression libraries and 150-bp paired-end reads for V(D)J libraries. Raw base call (BCL) files were converted to FASTQ format using Illumina software.

Single-cell RNA-seq reads were aligned to the GRCh38 human reference genome and quantified using Cell Ranger (version 8.0.1, 10X Genomics). To improve cell barcode recovery, droplets were additionally processed using the EmptyDrops algorithm implemented in the DropletUtils package (version 1.28.0), retaining both Cell Ranger- and EmptyDrops-identified cells. For sample demultiplexing, bi-allelic single-nucleotide polymorphisms (SNPs) were quantified at the single-cell level using cellsnp-lite (version 1.2.0), and donor identities were inferred using Vireo (version 0.5.8). Putative heterotypic doublets were identified using scDblFinder (version 1.22.0), while additional genotypic doublets were detected via Vireo. Ambient RNA contamination was corrected using SoupX (version 1.6.2). Genes detected in fewer than 10 cells were excluded from downstream analyses. Cell-level quality control criteria included (i) a mitochondrial transcript fraction <20% and (ii) detection of >300 genes per cell. Doublets identified by either scDblFinder or Vireo were removed prior to clustering and annotation.

Following preprocessing and initial annotation, 220,060 high-quality CD8+ T cells were selected from 195 samples across 120 donors (12 HG-T and 108 Non-T individuals), in which 150 samples were paired across pre- and post-treatment timepoints. Downstream analyses were performed using Scanpy (version 1.11.5), including log-normalisation and identification of top 3,000 highly variable genes (HVGs) within the CD8+ T population. To determine the optimal integration strategy, we benchmarked five widely used methods - BBKNN, Harmony, Scanorama, ComBat, and scVI - using the scIB-metrics framework (version 0.5.7). Based on batch correction scores and overall performance, scVI (version 1.1.5) was selected for batch correction, modelling each experimental pool as a batch covariate. Dimensionality reduction and clustering were performed using the scVI latent space, followed by nested Leiden clustering and visualisation using UMAP. For automated cell type annotation, scANVI (version 1.1.5) and CellTypist (version 1.7.1) were applied for cross-dataset label transfer using external references derived from Azimuth and COMBAT. In addition, the expression of canonical marker genes was also visualised to refine CD8+ T-cell cluster annotations.

### Subset-specific differential expression analysis with downsampling

Differential gene expression analysis was performed using Seurat (version 5.4.0) with the Wilcoxon rank-sum test implemented in the *FindMarkers()* function, unless otherwise specified. For longitudinal comparisons, only pre- and post-treatment paired samples with more than 20 cells per subset were included in the analysis to ensure sufficient statistical power and representation. To control for differences in statistical power arising from unequal cell numbers across groups, we implemented a subsampling strategy whereby each group was downsampled to the size of the smallest group (n = 1,200 cells). This procedure was repeated 100 times, and results were aggregated to mitigate sampling variability and reduce bias. As an orthogonal validation, DE was additionally assessed using a donor-aware linear mixed-effects model. Genes with non-zero variance in the subsampled dataset were included, and donor identity was modelled as a random effect to account for inter-individual variability. For post-versus pre-treatment comparisons, six donors per group were randomly selected to ensure balanced representation across conditions.

Pathway enrichment analysis was performed using clusterProfiler (version 4.18.4) with GOBP gene sets. Up- and down-regulated genes were analysed separately using a hypergeometric test, with the background defined as all detected genes. To compare pathway-level responses to immune checkpoint blockade (ICB) between HG-T and Non-T individuals across cell subsets, enrichment analyses were conducted on each subsampled dataset (n = 1,200 cells per group). Pathway-level differences were quantified as the delta frequency of enriched pathways between HG-T and Non-T groups across bootstrap iterations. This approach was adopted to maximise statistical power for pathway detection, particularly in subsets with limited DE signals.

### Single-cell TCR repertoire

TCR chains were filtered based on productive rearrangements and chain identity (TRA or TRB). Cells were retained if they contained either (i) a single TCRβ chain, (ii) one TCRα paired with one TCRβ chain, or (iii) two TCRα chains paired with one TCRβ chain. Cells not meeting these criteria were excluded from downstream analyses. In addition, samples with fewer than 100 cells containing detectable TCR clonotypes were removed to ensure sufficient repertoire coverage. Clonotypes were defined based on the concatenated amino acid sequences of CDR3 regions present within each cell, with cells sharing identical CDR3 compositions assigned to the same clonotype. Clonal proportions were calculated as the number of cells belonging to each clonotype divided by the total number of cells per sample that passed the above filtering criteria.

In total, 165,040 cells from pre- and post-treatment paired samples across 75 patients had matched transcriptomic data. After quality control, 112,897 cells from 72 individuals retained high-confidence V(D)J sequencing information with valid chain configurations and were included in all downstream clonal analyses. For analyses of clonal phenotype, cells were grouped by clonotype, individual, and time point. The distribution of each clone across transcriptional subsets was quantified as the proportion of its constituent cells assigned to each subset. The subset with the highest proportion was designated as the dominant phenotype of the clone. Definitions of persistent, new and lost clones were applied as described in the MiXCR-based bulk TCR repertoire analysis section.

### Pathway scoring with UCell

Single-cell gene set activity was quantified using UCell (version 2.14.0), a rank-based scoring method robust to differences in sequencing depth and data sparsity. Gene sets were obtained from the MSigDB Hallmark collection. To ensure adequate representation, only gene sets with more than five overlapping genes detected in the dataset were retained for analysis. UCell scores were computed at the single-cell level based on gene expression rankings. To summarise pathway activity at the clonal level, cells were grouped by clonotype, and the median UCell score across all cells within each clone was calculated for each gene set.

### Inference of transcription factor regulons

Gene regulatory network analysis was performed using the pySCENIC pipeline (version 0.12.2b0). The raw single-cell count matrix was used as input to infer transcription factor (TF)-target interactions. Co-expression relationships between TFs and candidate target genes were first estimated using the *GRNBoost2* algorithm implemented in the *grn* step. TF-target pairs with importance scores >1 were retained for downstream analysis. Regulons were subsequently refined using the *cisTarget (ctx)* step, which integrates motif enrichment analysis to identify direct TF targets. Motif annotation was performed using the *motifs-v10nr_clust-nr.hgnc-m0.001-o0.0.tbl* database, and genomic rankings were derived from the *hg38_10kbp_up_10kbp_down_full_tx_v10_clust.genes_vs_motifs.rankings.feather* reference. Regulon activity at the single-cell level was quantified using AUCell, which computes enrichment scores based on the ranked expression of target genes. To assess cell type specificity, regulons were ranked within each CD8+ T-cell subset based on their regulon specificity scores (RSS).

### Genotyping

The Illumina Global Screening Array 24 version 3 (Illumina) was used for genotyping. For sample deconvolution of pooled single-cell data, nuclear autosomal genotypes were aligned to hg37 and assessed for call frequency > 0.9, SNP missingness < 3%, Minor Allele Frequency (MAF) > 1%, and Hardy–Weinberg Equilibrium (HWE) P > 10-20 using GenomeStudio (version 2.0.5, Illumina) and PLINK (version 1.9b6.24); samples with call rates < 97% were removed from further analysis. Upon initial quality control, imputation was performed using the Michigan Imputation Server with Haplotype Reference Consortium (HRC) version 1.1 Build hg19 as a reference panel, as well as Eagle (version 2.4) and Minimac4 (version 1.7.4) for phasing and imputation. SNPs were further screened for MAF > 4%, HWE P > 10−10 and r^2^ > 0.7 and lifted-over to hg38.

### Prediction of mitochondrial haplogroups

For mitochondrial haplogroup prediction, mtDNA sites were aligned to the revised Cambridge Reference Sequence (rCRS). Initial QC was performed on mitochondrial variants, retaining SNPs with call rates > 90% and MAF > 0.1%. Alleles were flipped or switched as required to guarantee alignment with rCRS orientation. With samples designated as male, mitochondrial imputation was then carried out using a GenBank mitochondrial reference panel^82^ within the IMPUTE2 (version 2.3.2) chromosome X framework. Post-imputation QC included filtering variants with an imputation INFO score > 0.7 and resolving reference/alternate allele discrepancies. HaploGrep3 (version 3.2.1) was used to assign mitochondrial haplogroups based on PhyloTree build 17.2, applying a heteroplasmy level threshold of 0.9.

### Seahorse T cell Persistence Assay

Paired pre- and post-treatment PBMC samples were selected by an independent investigator prior to experimentation. The principal experimentalist remained blinded to patient cancer type, treatment regimen, lymphocyte stability and mitochondrial haplogroup status until completion of data quality control and normalisation.

Cryopreserved PBMCs were rapidly thawed in pre-warmed R10 (RPMI-1640 supplemented with 10% fetal bovine serum) and rested for 4 hours at 37°C in a 5% CO_2_ incubator to allow metabolic recovery. Untouched T cells were isolated from PBMCs using negative selection (Pan T cell isolation kit, Miltenyi Biotec) according to the manufacturer’s protocol. T cells were stimulated for 16 hours on anti-CD3 (10μg/ml, BioLegend) coated plates in the presence of soluble anti-CD28 (5μg/ml, BioLegend) to mimic antigen exposure and early activation-induced metabolic remodelling.

Activated T cells were resuspended in Seahorse XF RPMI medium supplemented with 10mM glucose, 1mM sodium pyruvate and 2mM L-glutamine as per manufacturer instructions (Agilent Technologies). Seahorse cell culture microplates were pre-coated with Poly-D-lysine (10μg/ml) for 1 hour at room temperature to facilitate cell adherence. Activated T cells were plated at 1 x 10^5^ total cells per well, with the maximum feasible number of technical replicates acquired for each sample according to cell availability. Plates were centrifuged at 200 x g for 2 minutes, and wells were visually inspected by light microscopy to confirm uniform cell seeding. Oxygen consumption rate (OCR) and extracellular acidification rate (ECAR) measurements were measured using the Seahorse XF Pro Analyzer (Agilent Technologies) at baseline and following sequential injection of oligomycin (1.5μM), BAM15 (2.5μM) and rotenone / antimycin A (0.5μM each).

To minimize inter-plate batch effects, paired samples from the same patient were always analysed on the same plate together with a standardized inter-plate healthy donor control. Wells failing pre-defined quality control criteria (e.g. abnormal mitochondrial stress test profiles, injection failure, poor cell adherence) were excluded. OCR and ECAR values were normalized to 1 x 10^5^ viable cells, and scaled relative to the basal respiration of the inter-plate control.

### Statistical analysis

All statistical analyses were performed in R (version 4.5.3). Survival analyses were conducted using survival (version 3.8-3) and survminer (version 0.5.1), and data visualisation was performed with ggplot2 (version 4.0.1). Unless otherwise specified, Cox proportional hazards models and logistic regression analyses were adjusted for age, sex, CMV serostatus, and treatment type (sICB versus cICB). Initial comparisons were performed using univariate log-rank tests in the metastatic melanoma cohort and subsequently validated in a pan-cancer cohort. Correlation analyses were performed using Spearman’s rank correlation coefficient. Comparisons between paired and unpaired groups were conducted using the Wilcoxon signed-rank test and Wilcoxon rank-sum test, respectively. Odds ratios (ORs) were estimated using Fisher’s exact test, and adjusted ORs were derived from generalized linear models with a binomial distribution. To evaluate the mediating role of LSI in the relationship between HG-T and OS, mediation analysis was performed using the mediation R package (version 4.5.1). A parametric Weibull survival model was used for the outcome model, as it provides a fully parametric framework that facilitates estimation of direct and indirect effects under time-to-event outcomes. Linear mixed-effects models were implemented in lme4 (version 1.1-38). All statistical tests were two-sided. Significance levels were defined as follows: P < 0.05, P < 0.01, P < 1×10^-3^, and P < 1×10^-4^. Where applicable, multiple testing correction was performed using the Benjamini-Hochberg (BH) method.

## Data Availability

All data produced in the present study are available upon reasonable request to the authors

## Acknowledgement

We thank all patients who contributed samples and participated in the study without compensation. We thank all the staff of the Day Treatment Unit, Oxford Cancer Centre and the Brodey Centre at the Horton General Hospital. We thank all the staff of the Oxford University Hospitals NHS Foundation Trust hematology, microbiology and biochemistry laboratories and thank T. James for his facilitation, as well as the staff of the Oxford Radcliffe Biobank and Churchill Hospital Sample Handling Lab. B.P.F. is funded by a Wellcome Career Development Award (grant no. 226535/Z/22/Z) and the study was previously funded by a Wellcome Intermediate Clinical Fellowship to B.P.F. (grant no. 201488/Z/16/Z). P.K. is funded by a Wellcome Discovery Award grant no. 222426/Z/21/Z and P.K. and B.P.F. are recipients of a CRUK Programme grant no. DRCNPG-Nov22/100005 ‘Defining the role of MAIT cells in Cancer Immunotherapy’. S.S. is a DPhil student funded by the Clarendon Fund, Department of Oncology Studentship and Jason Hu Scholarship from Balliol College Oxford. M.C. is a DPhil student funded by the Esther Yewpick Lee Millennium Scholarship. R.A.W. is funded by the CRIS cancer foundation post-doctoral fellowship (grant no. BBD00040), and previously by a Wellcome Trust Doctoral Training Fellowship (grant no. BST00070). G.M. is a DPhil student whose fees have been covered by Etcembly. M.L. is a DPhil student who is supported by Cancer Research UK (CRUK) (grant no. SEBCATP-2023/100003). The Oxford Radcliffe Biobank and Oxford Centre for Histopathology Research are supported by the University of Oxford, the Oxford CRUK Cancer Centre and the NIHR Oxford Biomedical Research Centre (Molecular Diagnostics Theme/Multimodal Pathology Subtheme), and the NIHR Cancer Research Network Thames Valley network. M.R.M. and B.P.F. are supported by the NIHR Oxford Biomedical Research Centre.

## Author Contributions

Analysis was led by S.S., M.C., R.A.W. and B.P.F.. M.C., S.M., and A.F. performed Seahorse functional assays. S.S. performed array genotyping imputation, variant-aware realignment and haplogroup prediction. S.S., M.C., and B.P.F. related lymphocyte stability to mitochondrial haplogroup. The sc dataset was developed by S.M., R.A.W., G.M., A.F., O.T., and D.M. with input from all authors, and the sc atlas was developed by S.S.. Sequencing QC and mapping of transcripts adaptive repertoires was performed by S.M., G.N., L.RZ.C. and S.S. M.L., B.P.F., M.J.P. and N.C. curated clinical data. S.S., M.C., R.A.W. and B.P.F. wrote the original draft of the manuscript and performed visualisation. B.P.F., M.L., M.R.M. and R.A.W. participated in project administration. B.P.F. and P.K. acquired funding. S.S. and M.C. contributed equally to this work. B.P.F. conceived and oversaw the study.

## Lead contact

Further information and requests for resources and reagents should be directed to and will be fulfilled by the Lead Contact, Benjamin P. Fairfax.

## Supplemental Information

### Supplementary tables

**Table S1** Clinical metadata of the OxCITE cohort (n = 417).

**Table S2** Clinical metadata and C2/C1 lymphocyte ratios of the external validation melanoma cohort (n = 170).

**Table S3** Differentially expressed genes (C2 vs. C1) in LS-S and LS-U patients across 100 bootstrap iterations, with the number of significant bootstraps and median log2 fold change.

**Table S4** Differentially expressed genes (C2 vs. C1) in LS-S and LS-U patients, analysed separately (DESeq2).

**Table S5** GOBP pathways significantly enriched in genes changing between C2 and C1 in LS-S and LS-U patients.

**Table S6** Seahorse functional measurements of OCR and ECAR at each time point in C1 and C2 samples, with derived metabolic parameters.

**Table S7** Differentially expressed genes (C2 vs. C1) in HG-T and Non-T patients, analysed separately.

**Table S8** GOBP pathways significantly enriched between C2 and C1 in HG-T and Non-T patients.

**Table S9** Differentially expressed genes between HG-T and non-T patients from the original alignment (DESeq2, adjusted for treatment cycle).

**Table S10** Baseline TPM-normalised expression of mtDNA-encoded protein-coding genes in bulk CD8+ T cells after NUMT remapping.

**Table S11** Nuclear-encoded genes used to score mitochondrial respiratory complexes I-V.

**Table S12** Proportions of CD8+ T cell subsets by patient and treatment cycle from single-cell RNA sequencing.

**Table S13** Number of subset-specific differentially expressed genes (C2 vs. C1) in HG-T and Non-T patients across 100 bootstrap iterations.

**Table S14** Number of significant bootstrap iterations for subset-specific GOBP pathways in HG-T and Non-T patients.

**Table S15** Sample availability for each OxCITE patient across the analyses and figures.

---

**Figure S1:**
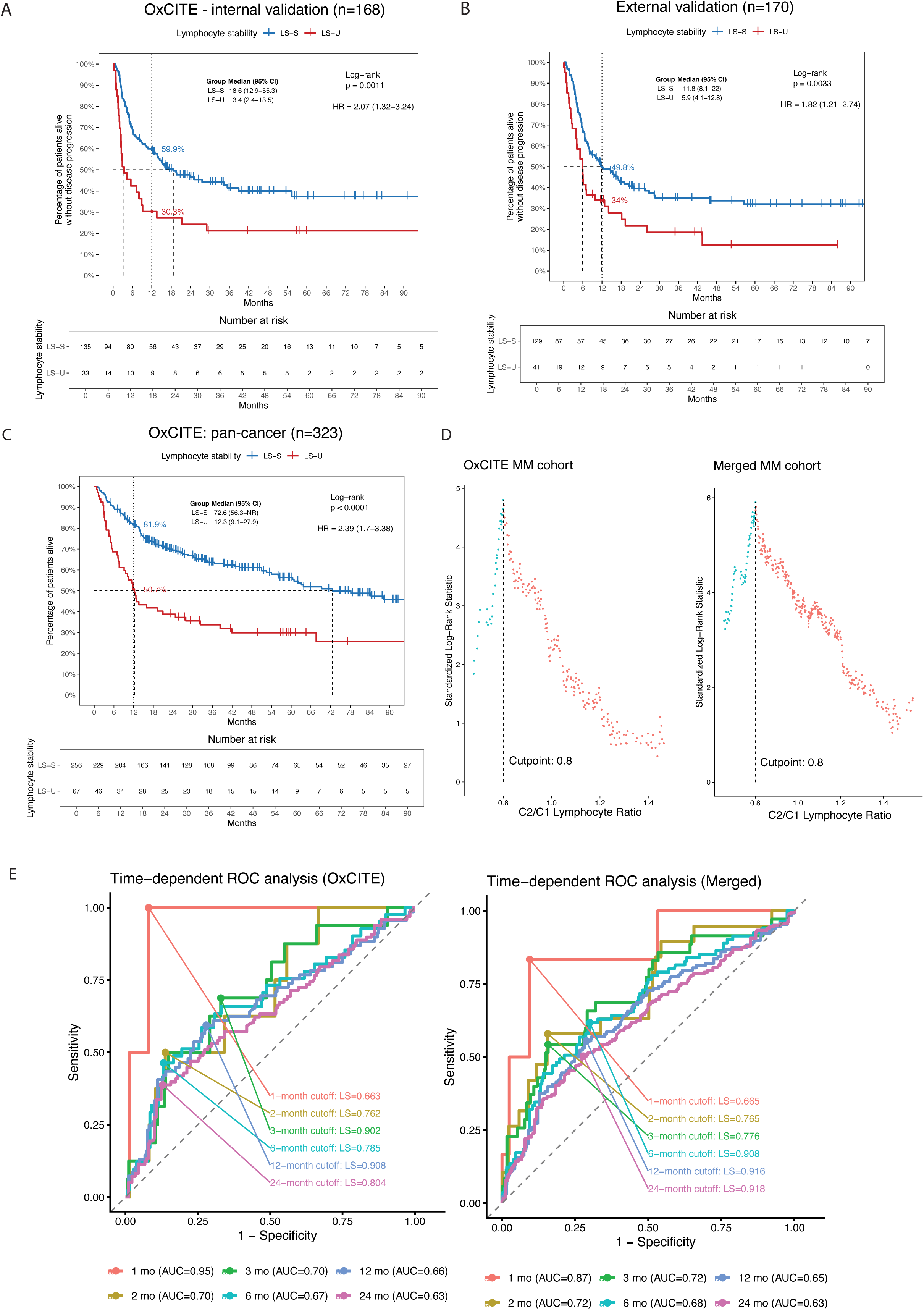
Cohort structure and validation analyses supporting lymphocyte stability (LS) as a biomarker. **(A)** Kaplan-Meier analysis of progression-free survival in the OxCITE internal validation melanoma cohort (n=168). **(B)** Kaplan-Meier analysis of progression-free survival in the external validation melanoma cohort (n=170). **(C)** Kaplan-Meier analysis of overall survival in the OxCITE pan-cancer cohort (n=323). **(D)** Standardized maximum log-rank statistic analysis supporting the C2/C1 lymphocyte ratio cutoff of 0.8 in the OxCITE melanoma and merged melanoma cohorts. **(E)** Time-dependent receiver operating characteristics (ROC) analyses across post-ICB timepoints in the OxCITE melanoma and merged melanoma cohorts, displaying time-specific LS cutoffs that yield maximal area under curve.

**Figure S2:**
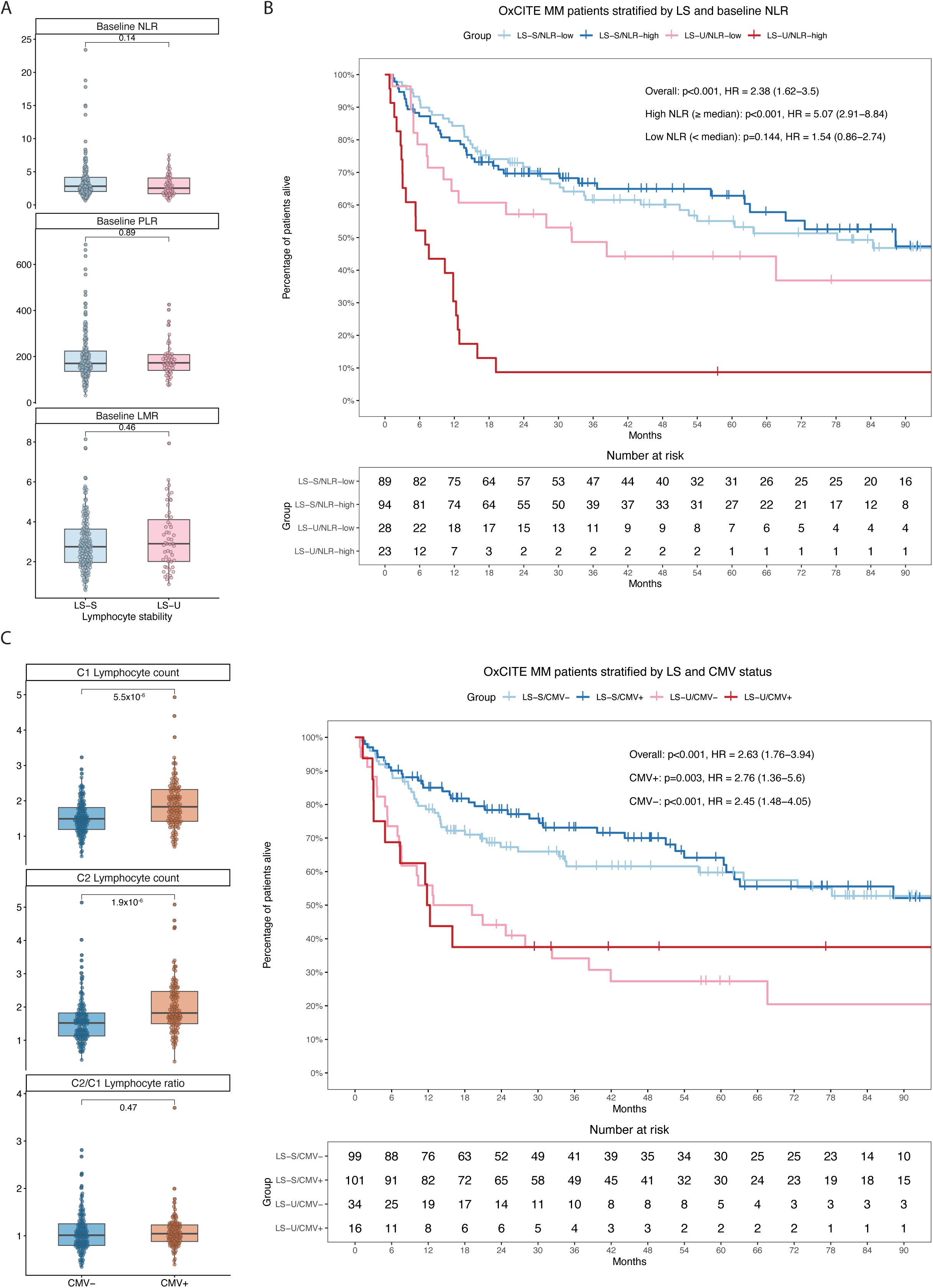
LS provides survival stratification beyond other blood-based indices and CMV serostatus. **(A)** Baseline neutrophil-to-lymphocyte ratio (NLR), platelet-to-lymphocyte ratio (PLR), and lymphocyte-to-monocyte ratio (LMR) in LS-S and LS-U patients. **(B)** Kaplan-Meier analysis of overall survival in OxCITE melanoma patients stratified by LS status and baseline NLR above or below the cohort median. **(C)** C1 lymphocyte count, C2 lymphocyte count, and C2/C1 lymphocyte ratio in CMV-seronegative and CMV-seropositive patients. **(D)** Kaplan-Meier analysis of overall survival in OxCITE melanoma patients stratified by LS status and CMV serostatus.

**Figure S3:**
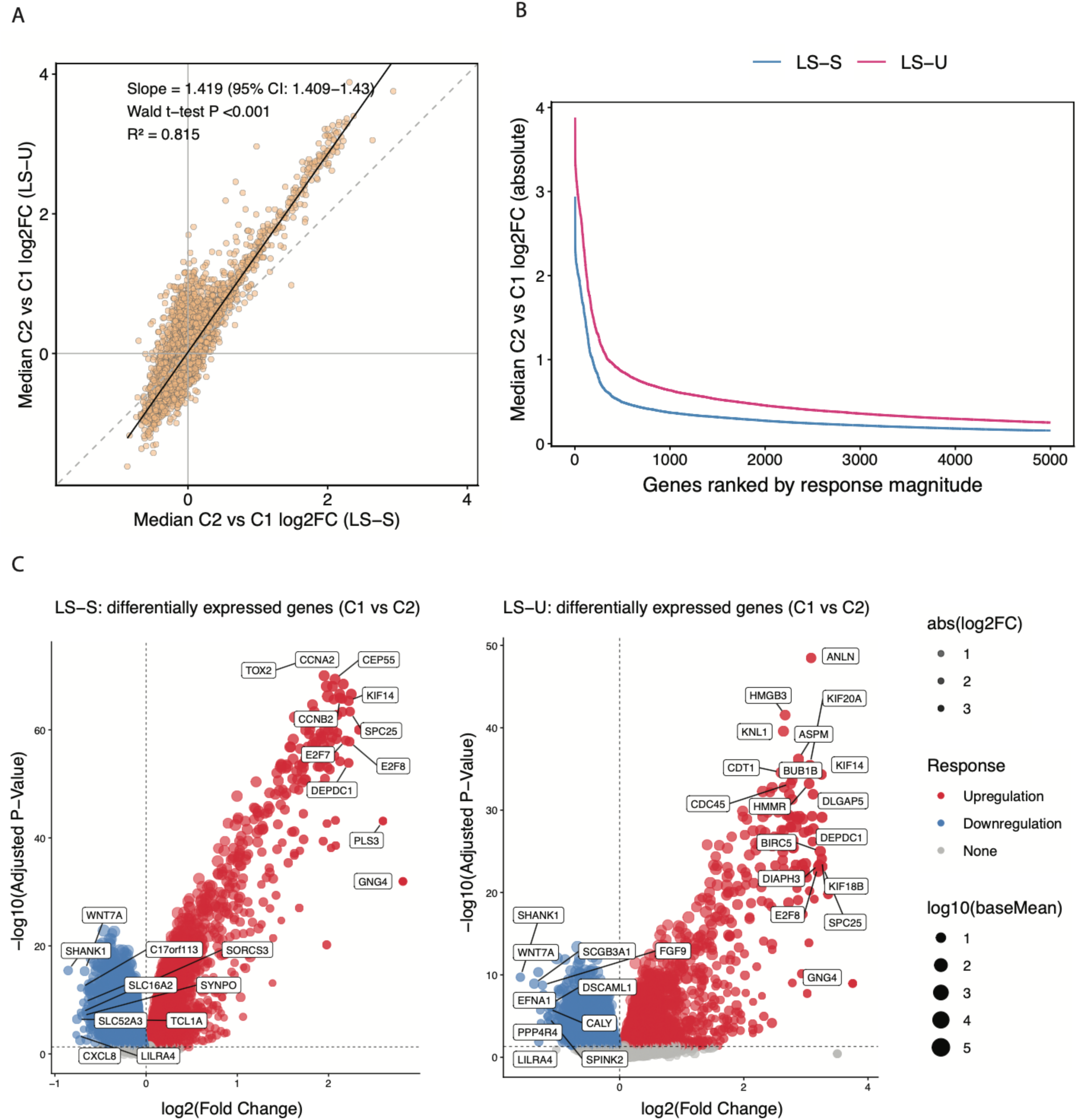
Supplementary analyses of ICB-induced CD8+ T-cell transcriptional remodelling. **(A)** Comparison of median ICB-induced gene-level log2FC in LS-U versus LS-S CD8+ T cells across 100 subsampling iterations (10 patients sampled from each LS group per iteration). Linear regression fit (solid line) with corresponding slope, Wald t-test P value and R² values shown. **(B)** Absolute median ICB-induced log₂ fold changes for genes ranked by response magnitude in LS-S and LS-U CD8+ T cells, showing greater transcriptional response magnitude in LS-U across the top 5,000 response genes., showing amplified transcriptional remodelling in LS-U. **(C)** Volcano plots of DEGs (C2 versus C1) in LS-S (left) and LS-U (right) CD8+ T cells.

**Figure S4:**
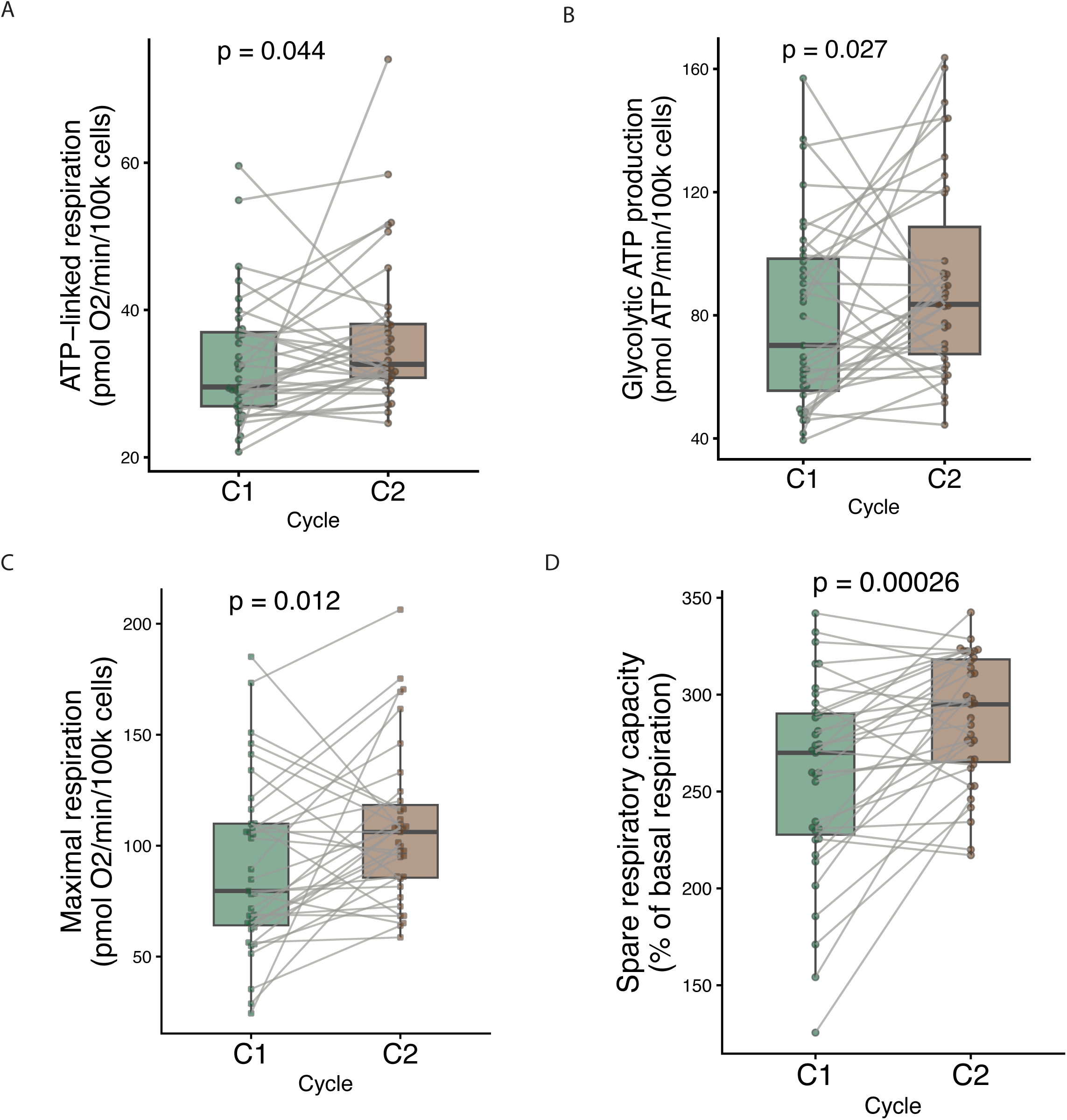
Supplementary analyses of T cell bioenergetic remodelling following ICB. **(A)** Paired ATP-linked respiration in T cells before and after ICB. **(B)** Paired glycolytic ATP production rate in T cells before and after ICB. **(C)** Paired maximal respiration in T cells before and after ICB. **(D)** Paired spare respiratory capacity (as a percentage of basal respiration rate) in T cells before and after ICB.

**Figure S5:**
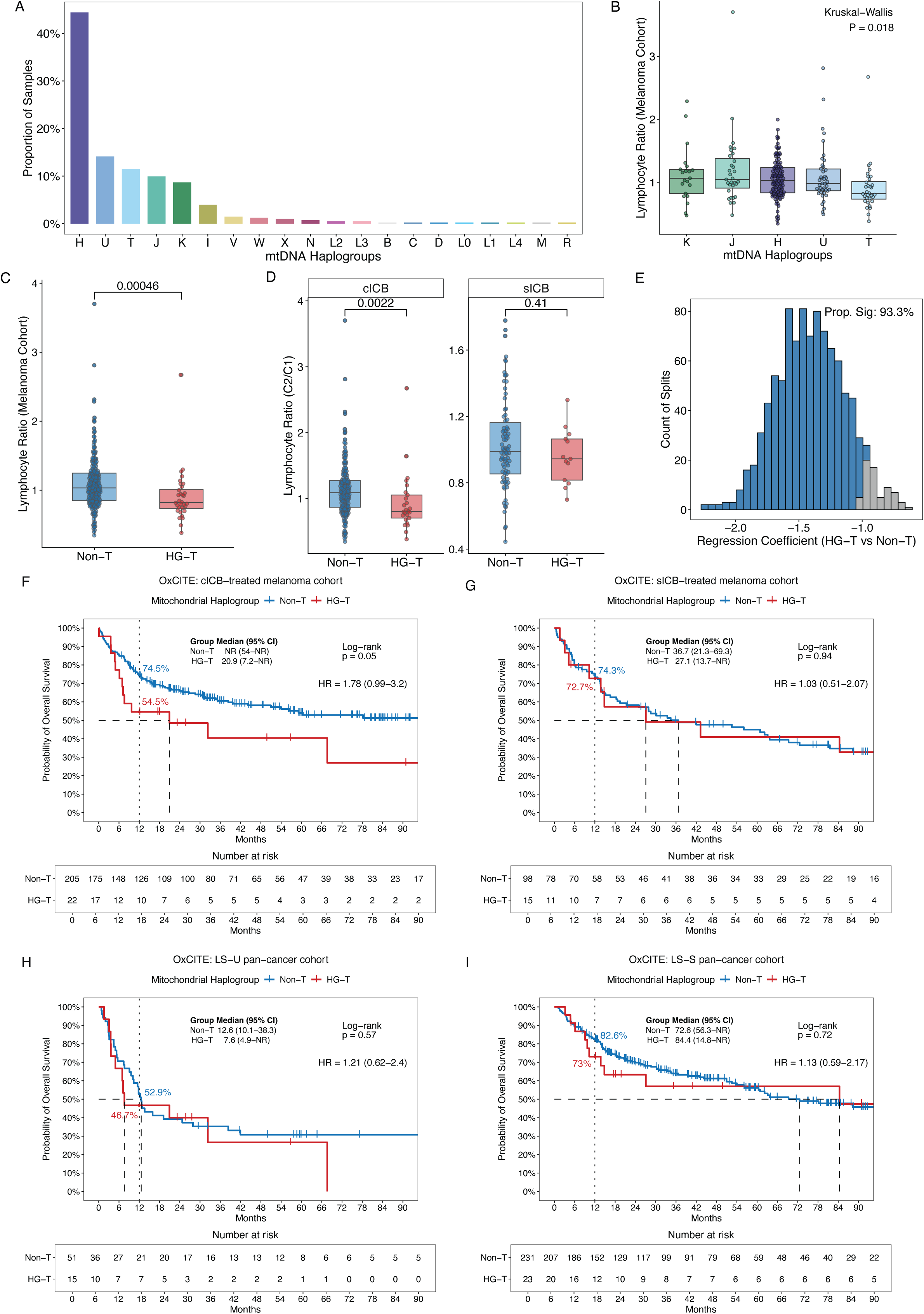
Mitochondrial HG-T is associated with lymphocyte ratio and ICB outcomes. **(A)** Distribution of mitochondrial haplogroups in the OxCITE cohort. **(B)** Lymphocyte ratio across major mitochondrial haplogroups in the OxCITE melanoma cohort. Differences were assessed using the Kruskal–Wallis test. **(C)** Lymphocyte ratio in HG–T versus Non–T patients in the OxCITE melanoma cohort. **(D)** Lymphocyte ratio in HG–T versus Non–T patients stratified by cICB and sICB treatment. **(E)** Bootstrap distribution of the regression coefficients for HG–T versus Non–T tests. The proportion of bootstrap estimates supporting the observed direction of association is indicated. **(F)** Kaplan-Meier analysis of OS by T haplogroup status in the cICB-treated melanoma cohort. **(G)** Kaplan-Meier analysis of OS by T haplogroup status in the sICB-treated melanoma cohort. **(H)** Kaplan-Meier analysis of OS by T haplogroup status in the LS-U pan-cancer cohort. **(I)** Kaplan-Meier analysis of OS by T haplogroup status in the LS-S pan-cancer cohort.

**Figure S6:**
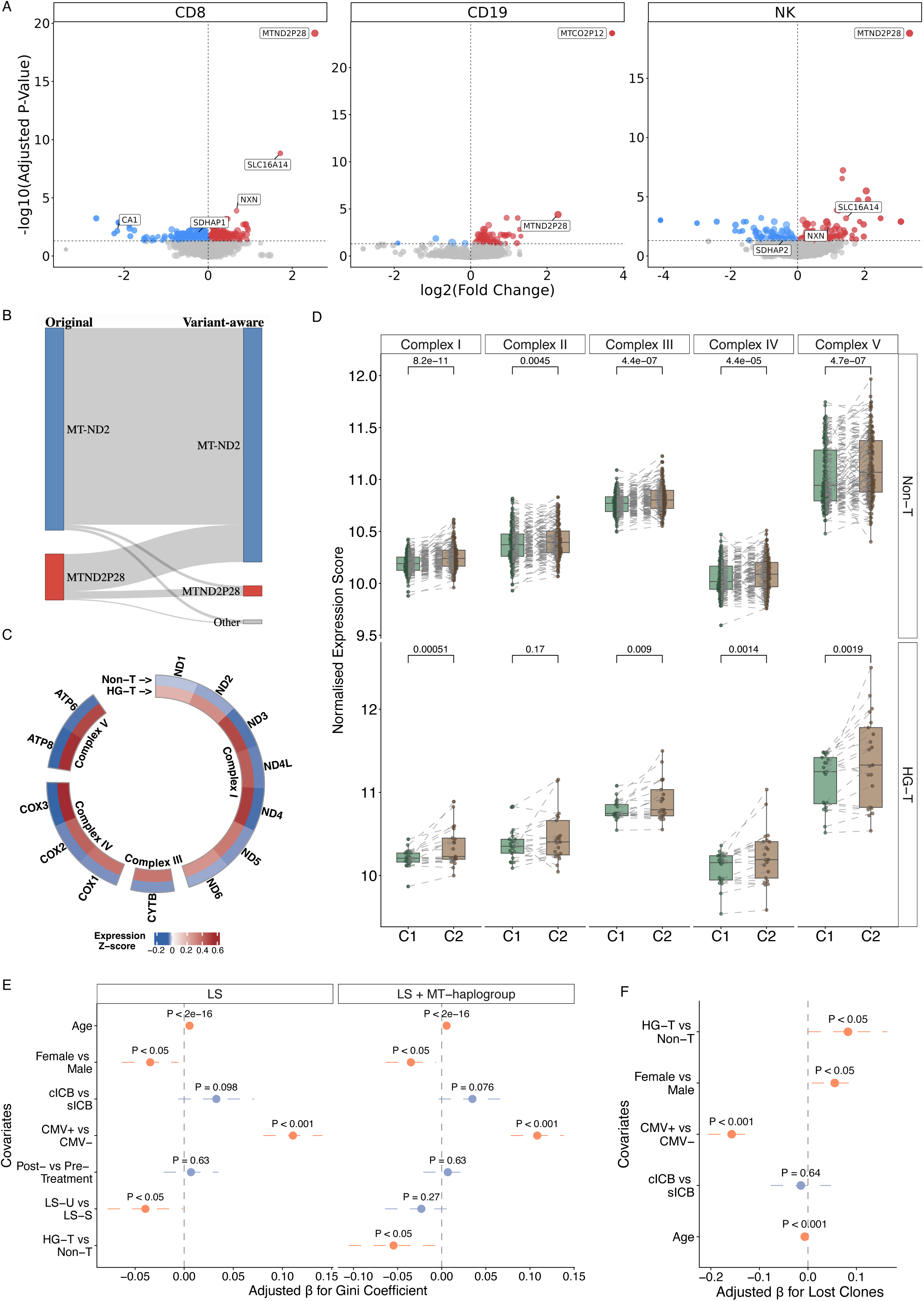
Distinctive HG-T transcriptional programs and clonal architecture of CD8+ T cells during checkpoint immunotherapy. **(A)** Differential gene expression between post- and pre-treatment in CD8+ T, CD19+ B, and NK cells. Top-ranked significant DE genes (adjusted P < 0.05) are labelled. **(B)** Aggregate fragment-level reassignment between mitochondrial *MT-ND2* and the homologous NUMT *MTND2P28* following mitochondrial variant-aware realignment. Fragments assigned to either locus in the original alignment were tracked following realignment to the variant-aware GRCh38 reference. Flow widths represent absolute fragment numbers; assignments outside either locus are grouped as “Other”. **(C)** Circular heatmap of baseline expression of the 13 mtDNA-encoded protein-coding genes in HG-T and Non-T CD8+ T cells, grouped by respiratory-chain complex. Colours represent group-mean gene-wise Z-scores of log2(TPM + 1) expression standardized across baseline samples. Outer and inner rings represent Non-T and HG-T, respectively. **(D)** Paired comparison of mitochondrial respiratory chain complex expression between C1 and C2, stratified by HG-T status. The expression scores are calculated as geometric means of complex-specific nuclear-encoded gene expression values. Lines connect paired samples. **(E)** Multivariable linear regression of factors associated with Gini coefficient, with or without adjustment for mitochondrial HG-T. Adjusted β coefficients and 95% CIs are shown. **(F)** Multivariable analysis of factors associated with lost T-cell clones.

**Figure S7:**
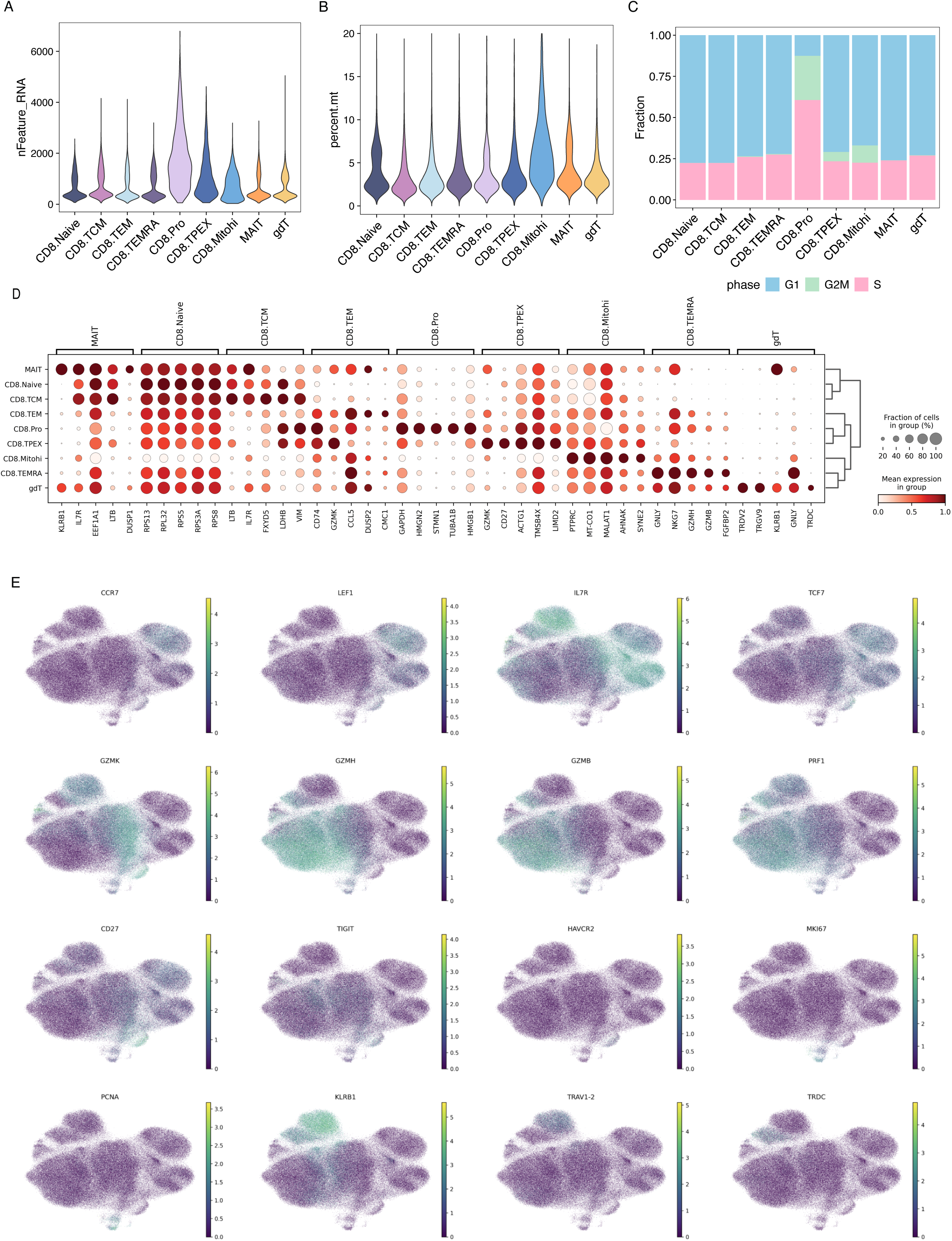
Single-cell atlas of peripheral CD8+ T-cell subsets before and after ICB. **(A)** Number of detected genes per cell across CD8+ T cell subsets. **(B)** Percentage of mitochondrial transcripts across CD8+ T cell subsets. **(C)** Cell-cycle phase composition across CD8+ T cell subsets. **(D)** Expression of cluster-specific marker genes across CD8+ T cell subsets. Dot size indicates the fraction of cells expressing each gene, while colour indicates mean expression. **(E)** UMAP visualisation of selected marker genes defining CD8+ T cell states.

**Figure S8:**
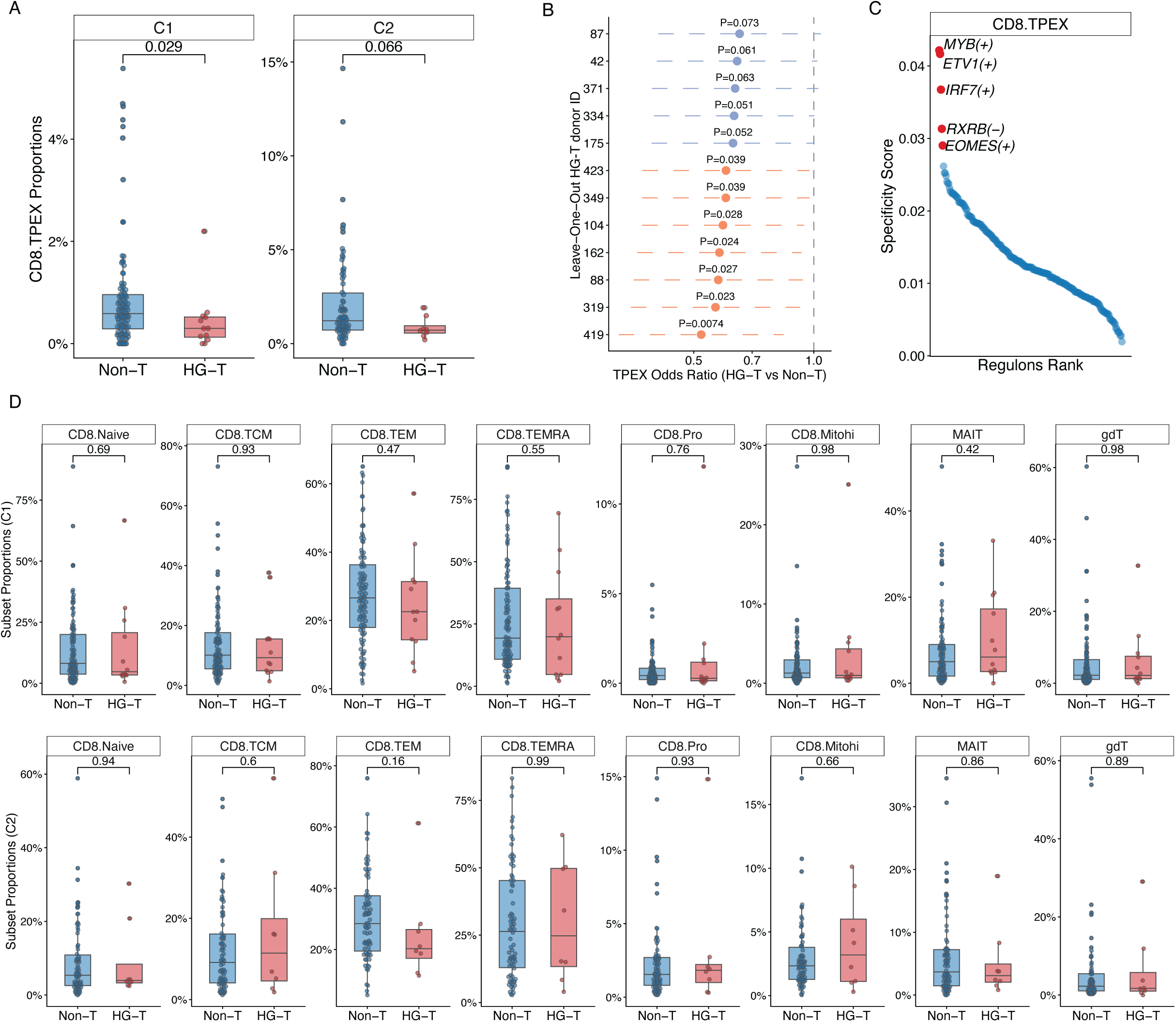
Dynamic remodelling of immune cell compositions in HG-T following ICB. **(A)** Proportions of CD8+ TPEX cells in HG-T and Non-T patients at C1 and C2. **(B)** Association between T haplogroup status and CD8+ TPEX abundance in leave-one-out analyses. ORs and 95% CIs are shown. **(C)** Regulon specificity ranking in CD8+ TPEX cells. Top-ranked transcription factor regulons are highlighted. **(D)** Proportions of other CD8+ T cell subsets in HG-T and Non-T patients at C1 and C2.

**Figure S9:**
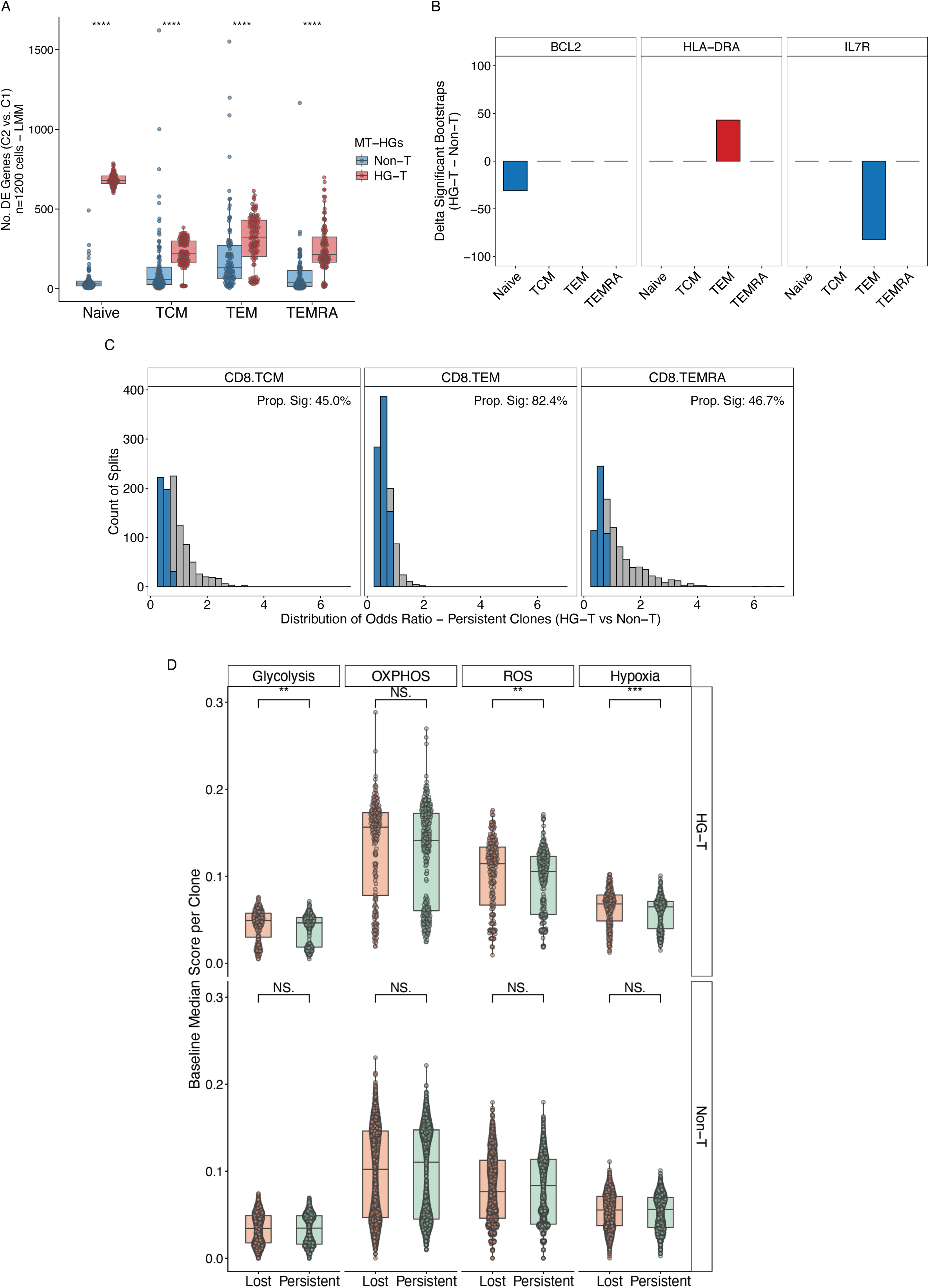
Subset-specific transcriptional and clonal signatures of CD8+ T cells in HG-T. **(A)** Numbers of DE genes between C2 and C1 identified by bootstrap-based linear mixed model across CD8+ T cell states in HG-T and Non-T patients. **(B)** Differences in the number of significantly enriched bootstrap iterations for selected genes between HG-T and Non-T groups across CD8+ T cell states. **(C)** Bootstrap distributions of ORs for persistent clones comparing HG-T with Non-T patients across CD8+ TCM, TEM, and TEMRA cells. The proportion of significant bootstrap iterations is indicated. **(D)** Baseline glycolysis, oxidative phosphorylation (OXPHOS), reactive oxygen species (ROS), and hypoxia scores of lost and persistent T cell clones, stratified by HG-T status.

**Extended Figure 1:**
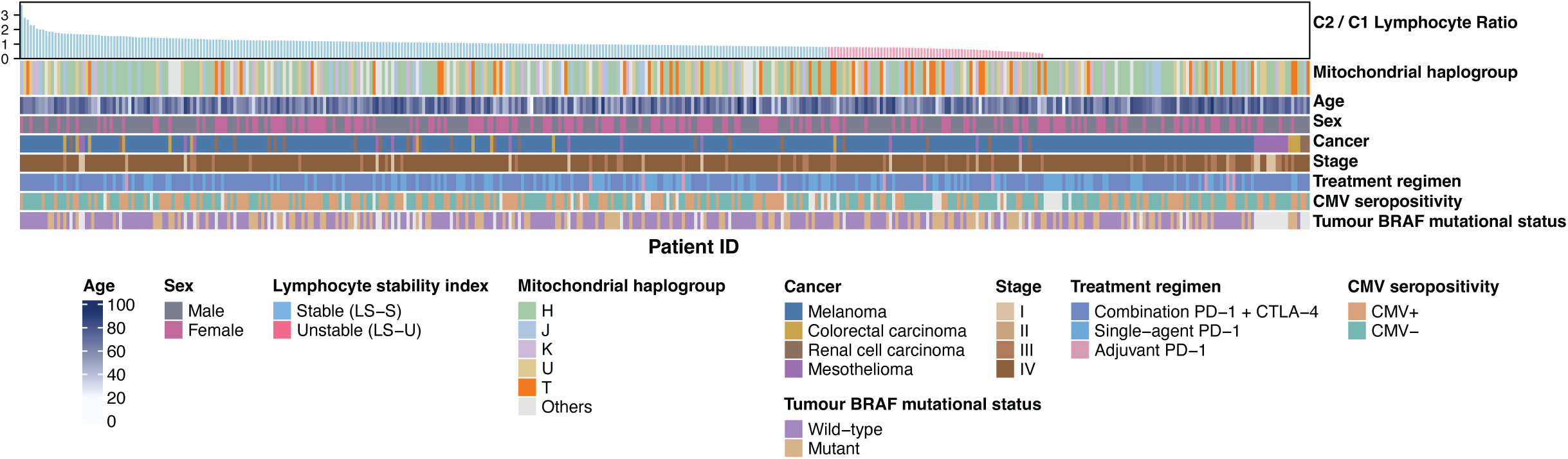
Baseline clinical characteristics of the OxCITE cohort stratified by LS status. Characteristics of patients treated with immune checkpoint blockade in the OxCITE cohort (n = 417), shown overall and for LS-stable (LS-S, n = 261) and LS-unstable (LS-U, n = 70) patients.

**Extended Table 1:** Clinical and genetic landscape of the OxCITE cohort. Each column represents one patient (n = 417). Tracks show C2/C1 lymphocyte ratio, lymphocyte stability, mitochondrial haplogroup, age, sex, cancer type, stage, treatment regimen, CMV serostatus and tumour BRAF mutational status.

| **Characteristic** | **Overall**<br>N = 417<sup>1</sup> | **Stable (LS-S)**<br>N = 261<sup>1</sup> | **Unstable (LS-U)**<br>N = 70<sup>1</sup> | **P value**<sup>2</sup> |
|---|---|---|---|---|
| **Age** | 68 (18–96) | 67 (18–96) | 67 (23–88) | 0.677 |
| **Sex** |  |  |  | 0.219 |
| Male | 245 (59%) | 148 (57%) | 46 (66%) |  |
| Female | 172 (41%) | 113 (43%) | 24 (34%) |  |
| **Cancer** |  |  |  | 0.876 |
| Melanoma | 353 (85%) | 223 (85%) | 62 (89%) |  |
| Mesothelioma | 25 (6.0%) | 11 (4.2%) | 3 (4.3%) |  |
| Colorectal carcinoma | 15 (3.6%) | 10 (3.8%) | 1 (1.4%) |  |
| Renal cell carcinoma | 24 (5.8%) | 17 (6.5%) | 4 (5.7%) |  |
| **Stage** |  |  |  | 0.528 |
| I | 15 (3.6%) | 8 (3.1%) | 2 (2.9%) |  |
| II | 4 (1.0%) | 2 (0.8%) | 1 (1.4%) |  |
| III | 43 (10%) | 20 (7.7%) | 8 (11%) |  |
| IV | 355 (85%) | 231 (89%) | 59 (84%) |  |
| **Treatment regimen** |  |  |  | 0.381 |
| Combination PD-1 + CTLA-4 | 279 (67%) | 177 (68%) | 49 (70%) |  |
| Single-agent PD-1 | 127 (30%) | 79 (30%) | 18 (26%) |  |
| Adjuvant PD-1 | 11 (2.6%) | 5 (1.9%) | 3 (4.3%) |  |
| **CMV seropositivity** |  |  |  | 0.010 |
| CMV+ | 172 (45%) | 121 (50%) | 19 (31%) |  |
| CMV- | 211 (55%) | 122 (50%) | 42 (69%) |  |
| **Tumour BRAF mutational status** |  |  |  | 0.052 |
| Wild-type | 229 (63%) | 156 (67%) | 33 (53%) |  |
| Mutant | 133 (37%) | 76 (33%) | 29 (47%) |  |
| **Mitochondrial haplogroup** |  |  |  | 0.003 |
| HG-T | 49 (12%) | 23 (8.8%) | 16 (23%) |  |
| Non-T | 367 (88%) | 238 (91%) | 53 (77%) |  |

<sup>*1*</sup> Median (Min–Max); n (%)
<sup>*2*</sup> Wilcoxon rank sum test; Fisher’s exact test

